# Development and validation of a district-level index of socioeconomic development for India

**DOI:** 10.1101/2025.04.02.25325109

**Authors:** Vinita Subramanya, Shivani A. Patel, Shakira F. Suglia, Robin A. Richardson, K.M. Venkat Narayan, Alvaro Alonso

## Abstract

**Introduction:** Socioeconomic development influences population health outcomes. The coexistence of rapid economic growth and persistent health inequalities in India underscores the need for community-level assessments of socioeconomic development. We created and validated four distinct multidimensional indices of socioeconomic development at the district level in India using separate data sources and two different methods.

**Methods:** Using data from the 2011 Census of India and the National Family Health Survey-4 (NFHS-4), four indices were constructed through principal components analysis (PCA) and percentile ranking. These indices covered sociodemographic, socioeconomic, housing, and community infrastructure domains. The indices were assessed for construct and criterion validity, examining their ability to capture the latent constructs of socioeconomic development and their relationship with health outcomes.

**Results:** All four indices suggested geographic disparities in socioeconomic development across India, with development lower in central regions and higher in southern states. They all demonstrated good construct validity, particularly with NFHS-4 data. The indices exhibited strong internal and external concurrent validity, aligning with the Multidimensional Poverty Index while also demonstrating predictive validity for nutritional health outcomes. Negative correlations with undernutrition in children and women and a positive correlation with being overweight or obese in adults were observed.

**Conclusion:** We successfully developed and validated multidimensional indices to measure socioeconomic development at the district level in India. Using both the 2011 Census and NFHS-4 ensured a comprehensive, multidimensional perspective on socioeconomic development and its health implications. The combination of PCA (for data-driven index creation) and percentile ranking (for relative comparisons) enhanced methodological robustness, providing a validated, scalable approach for assessing district-level development and its impact on public health in India. Future temporal and spatial analyses should explore the causal relationship between socioeconomic development and health outcomes.

## Introduction

Area-level social and material factors, including access to nutritious food, safe spaces for physical activity, healthcare, sanitation, and exposure to pollutants, are influenced by socioeconomic development, impact health behaviors, contributing to area-level health disparities.^1–4^ Individuals in socioeconomically disadvantaged communities experience disproportionate adverse health outcomes even after accounting for individual socioeconomic characteristics. ^1,2,5–10^

India, middle income country (World Bank classification), presents a complex interplay of rapid economic growth and health disparities. Early research in India predominantly relied on unidimensional poverty measures; however, contemporary studies have shifted toward multidimensional frameworks that better capture the complexity of economic development.

Recognizing the substantial influence of community-level conditions on health outcomes, these frameworks consider compositional, contextual, and collective factors, drawing conceptually from Peter Townsend’s economic deprivation theory and Amartya Sen and Mahbub-ul-Haq’s development theories. ^11–17^

Building on this, a substantial body of work in India has focused on developing socioeconomic indices that incorporate housing conditions, asset ownership, income and broader health and infrastructure indicators. Methods for creating these indices are varied and may employ arithmetic or geometric means, principal components analysis, the Wroclaw Taxonomic Method, and the Alkire-Foster method.^18–22^ These indices assess geographical disparities in socioeconomic development and its impact on health, revealing patterns such as lower utilization of maternal health services and higher COVID-19 incidence in disadvantaged areas.^23–25^ Prior studies have not systematically evaluated publicly available datasets to determine which might be more effective in developing an index of socioeconomic development, nor have they compared different methodological approaches to assess their relative strengths and limitations. Additionally, few indices have been validated to ensure they accurately capture intended constructs across different subpopulations and contexts, a process complicated by the lack of a gold standard for validation.^26^

Research on the relationship between socioeconomic development and health in India, especially chronic conditions like cardiovascular disease, remains limited. A conceptually grounded and validated index of community-level socioeconomic development is needed to evaluate health outcomes and track determinants of health trajectories, vulnerable populations, and policy intervention targets.

We developed and validated contextually relevant indices of socioeconomic development to understand and monitor health inequalities in India. Using two national datasets (Census 2011 and NFHS-4) and two methodological approaches to develop four distinct indices, we created four distinct indices, enabling independent validation of socioeconomic constructs and enhancing the robustness of our findings. By employing both PCA and percentile ranking separately, we undertook a comparative assessment of which approach best predicts health outcomes. Our objectives were to (i) construct district-level socioeconomic development indices using two methods and datasets and (ii) validate these indices through construct and criterion validity assessments.

The objectives were to i) construct socioeconomic development indices at the district level using two methods and datasets and ii) validate the indices by assessing construct and criterion validity.

## Methods

Two publicly existing datasets were analyzed using two separate methodologies to construct and validate indices of socioeconomic development. **Figure 1** and **Supplemental Table 1** present an overview of the data sources and methods. Specifically, we drew on data from the Census of India (2011) and the National Family Health Survey-4 (NFHS-4) available on the National Data Analytic Platform (NDAP), to construct four indices using principal components analysis and percentile ranking, respectively. For each index, indicators were selected across four domains. All four indices were evaluated for construct and criterion validity. More details on the NDAP platform are in the **Supplemental Methods**.

**Figure 1:**
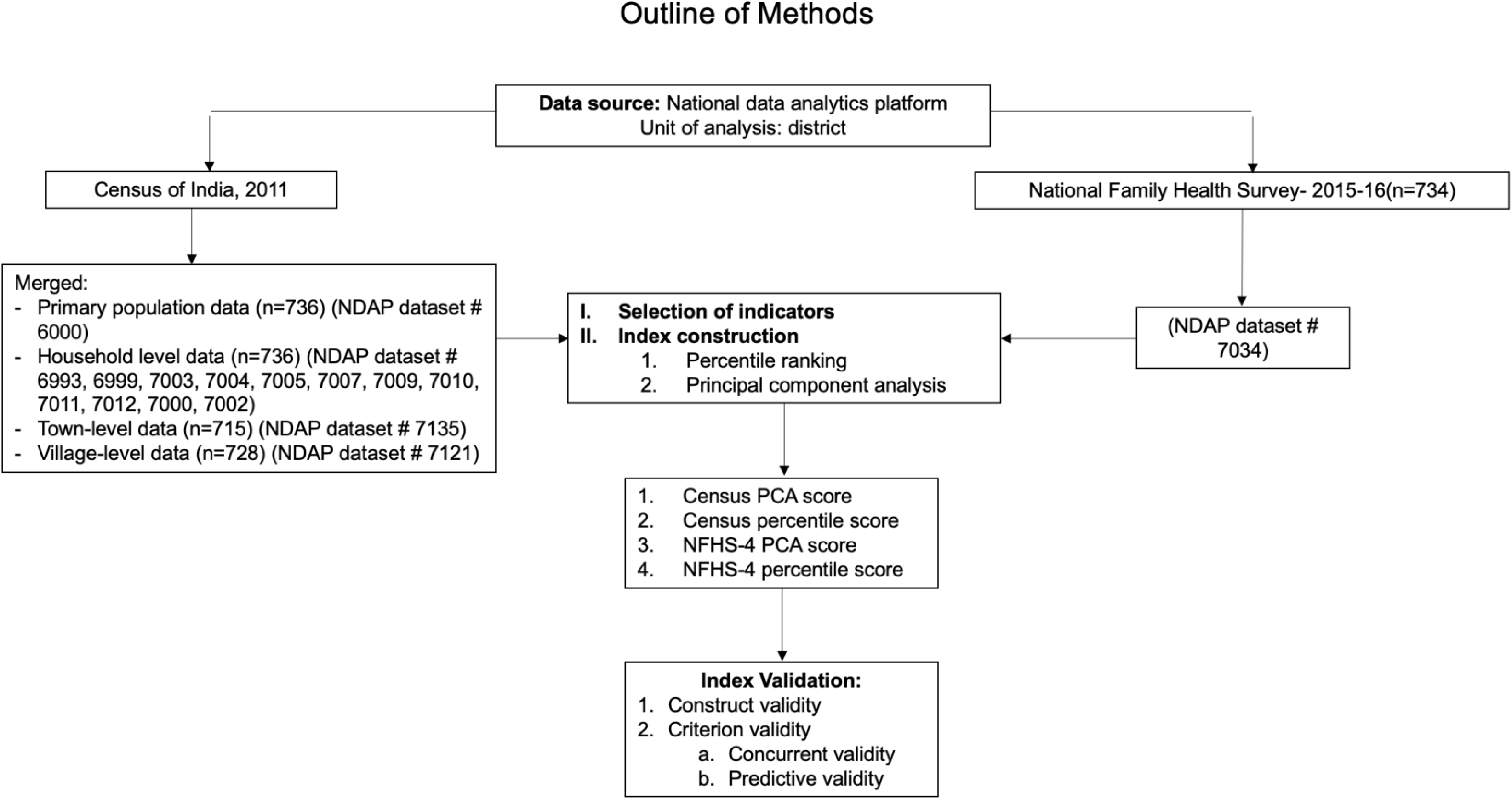
Outline of analytical methods used in primary index construction and validation

### Unit of analysis

India has 28 states and 8 union territories, subdivided into smaller administrative units for governance. Districts, headed by a district magistrate, serve as administrative sub-divisions. The district was chosen due to the availability of several datasets at this level of granularity. NDAP data was standardized to provide information on up to 765 districts.

### Datasets

#### a. Census of India 2011

The decennial Census enumerates the population and captures demographic and sociocultural information. The 2011 Census^27–30^ covered 28 states and 6 union territories, collecting data on housing stock and household amenities in Phase I, and detailed individual attributes in Phase II. Available Census data in NDAP varied from 736 districts (household-level), 728 (village-level), and 715 (town-level).

#### b. National Family Health Survey-4

The NFHS-4^31^ (2015-16) is a nationally representative survey assessing health, family welfare, and emerging issues at the individual level. It used a stratified two-stage sampling strategy, selecting primary sampling units and households.^31^ The survey is suitable to derive health estimates at national, state, union territory, and district levels. NFHS-4 data in NDAP covered 734 districts.

### Index development

#### i. Selection of Indicators

Indicators were chosen from four domains: Sociodemographic, Socioeconomic, Housing, and Community infrastructure. Variables were retained as such or transformed, as needed. The fifteen NFHS-4 and the thirty-five Census indicators used in index construction have been described below and listed in **Supplemental Table 2** and **Supplemental Figure 1**.

**Sociodemographic indicators** included population below 15 years, sex ratio at birth, and early marriage rates from NFHS-4, and population below 6 years, urban/rural populations, and scheduled caste/tribe data from the Census. **Socioeconomic indicators** included literacy rates and employment classification from the Census and literacy rates from NFHS-4. **Housing indicators in the Census and NFHS-4** covered household standard of living characteristics and assets.^37^ **Community infrastructure and resource indicators** included the number of schools from the Census and healthcare facilities and civil registration services from NFHS-4.

**Health outcomes** from NFHS-4 included nutritional indicators in children and adults and cardiovascular risk factors in adult men and women. Nutritional indicators included infant breastfeeding, childhood wasting and stunting, and anemia in children and adults. Cardiovascular risk factors included hypertension, diabetes mellitus and being overweight or obese.

#### ii. Index construction

Two methods were used: Principal Components Analysis (PCA) and percentile ranking. PCA is a dimension reduction technique used to summarize the variation contained in the original set of variables through a smaller set of continuous scores known as components. Under the PCA methodology, the index was based on the first component, representing the largest contribution of variance in the data.^57,58^ While PCA focuses on indicators that are most influential for variation (or separation) across data points, percentile ranking focuses on relative position of data points across observations. Percentile ranking is a method that was used to assign each unit district a percentile score ranging from 0 to 100 based on the rank of each indicator from lowest to highest. Under the percentile ranking method, the index was the sum of the percentile ranking across all indicators (a higher score indicates greater development).^33,59^ Both the PCA and percentile methods were applied to the two datasets to develop and evaluate a total of four overall district-level scores. The distribution of indicators by district, from each dataset, were described using the mean (standard deviation), median, range and interquartile range. Additionally, urban-rural stratified scores were developed (**Supplemental Methods**).

### Index validation

All indices were evaluated for construct validity and criterion validity.

i. Construct validity, the degree to which the index is related to its constituent indicators, as expected by proposed theoretical relationships, was evaluated by examining the correlations between indicators. ^26,60^ We assessed for convergent validity, where indicators in a given domain are related. Additionally, we evaluated correlations between the index and constituent indicators, i.e., the factor loadings in the PCA analysis.^60^
ii. Criterion validity is whether the index varies or relates to an external index or criteria. We evaluated for two types of criterion validity – concurrent and predictive validity. Without an established, widely used index, we compared Pearson correlations of the scores developed by different methods within the dataset for internal concurrent validity. ^26,60^ We also compared domain-specific scores across datasets. Strong positive correlations would suggest good concurrent validity. To evaluate external concurrent validity, we compared district-level maps of socioeconomic development between the overall scores of this analysis and the Multidimensional Poverty Index (**Supplemental Table 3**).^61^ The index scores aimed to assess area-level inequalities and their effects on nutritional indicators and cardiovascular risk factors. Therefore, Pearson correlations between each area-level socioeconomic development index and specified health outcomes measured in the NFHS-4 were assessed as a measure of predictive validity. ^26,60^

## Results

### I. Index construction

#### a. Distribution of indicators by district (**Table 1**)

##### Census

Approximately two-thirds of the Indian population resided in rural areas. Villages had more healthcare facilities (per 10,000 people) than towns, perhaps due to the large network of informal healthcare providers in villages. A majority of occupied census houses [median (interquartile range): 91% (83-96%) were owned. The median (IQR) percentage of homes with drinking water from a treated tap water source was 22% (10-41%), an improved latrine facility was 37% (22-59%), a bathroom on premises was 32% (16-58%), a kitchen on premises was 59% (40-78%), and 19% (9-35%) of homes used LPG as cooking fuel, suggesting a slow transition from solid or biomass fuels to cleaner fuel options.

**Table 1:** Population and community characteristics-National Family Health Survey-4, Census of India.

| National Family Health Survey- 4 (2015-16) |  |  |  |  |  |  |  |
| --- | --- | --- | --- | --- | --- | --- | --- |
| Variable | N | Mean | SD | p25 | p50 | p75 | IQR |
| Households using clean fuel for cooking (%) | 728 | 38 | 23 | 19 | 34 | 54 | 35 |
| Population living in households with an improved drinking-water source (%) | 728 | 87 | 14 | 82 | 92 | 97 | 15 |
| Population living in households that use an improved sanitation facility (%) | 728 | 48 | 22 | 31 | 47 | 65 | 35 |
| Population living in households with electricity (%) | 728 | 89 | 14 | 85 | 95 | 98 | 14 |
| Population below age 15 years (%) | 728 | 29 | 6 | 24 | 28 | 33 | 8 |
| Sex ratio at birth for children born in the last five years<sup>1</sup> | 728 | 936 | 115 | 862 | 930 | 1003 | 141 |
| Women in the age group of 20 to 24 years married before age 18 years (%) | 728 | 25 | 13 | 14 | 23 | 34 | 20 |
| Men age group 15 to 49 years who are literate (%) | 728 | 85 | 8 | 80 | 86 | 91 | 11 |
| Women age group 15 to 49 years who are literate (%) | 728 | 68 | 14 | 59 | 69 | 78 | 20 |
| Institutional births (%) | 728 | 78 | 18 | 68 | 83 | 93 | 25 |
| Births delivered by skilled health personnel (%) | 728 | 81 | 16 | 72 | 85 | 94 | 22 |
| Registered pregnancies (%) | 728 | 89 | 12 | 85 | 92 | 96 | 11 |
| Children under age 5 years whose birth was registered with the civil authority (%) | 728 | 82 | 16 | 69 | 86 | 96 | 27 |
| Households with any usual member covered under insurance scheme (%) | 728 | 29 | 24 | 10 | 20 | 48 | 38 |
| Mothers who received financial assistance under janani suraksha yojana (%) | 728 | 39 | 23 | 18 | 36 | 61 | 42 |
| Census of India 2011 |  |  |  |  |  |  |  |
| ***Town characteristics\**** |  |  |  |  |  |  |  |
| No. of latrines (pit and flush and pour) | 715 | 1466 | 640 | 1007 | 1448 | 1919 | 912 |
| No. of domestic electric connections | 715 | 1726 | 639 | 1291 | 1703 | 2087 | 796 |
| No. of industrial electric connections | 715 | 46 | 90 | 12 | 26 | 48 | 36 |
| No. of commercial electric connections | 715 | 308 | 324 | 164 | 263 | 401 | 237 |
| No. of electric connections for road lighting | 715 | 261 | 191 | 116 | 250 | 369 | 253 |
| No. of healthcare facilities | 715 | 2 | 3 | 1 | 2 | 3 | 2 |
| No. of primary schools (government and private) | 715 | 6 | 5 | 4 | 6 | 8 | 4 |
| No. of middle schools (government and private) | 715 | 4 | 4 | 2 | 3 | 5 | 3 |
| No. of secondary schools (government and private) | 715 | 2 | 1 | 1 | 2 | 3 | 2 |
| No. of senior secondary schools (government and private) | 715 | 1 | 1 | 1 | 1 | 2 | 1 |
| ***Village characteristics\*\**** |  |  |  |  |  |  |  |
| No. of primary schools (government and private) | 728 | 12 | 7 | 8 | 11 | 15 | 8 |
| No. of middle schools (government and private) | 728 | 5 | 3 | 3 | 5 | 7 | 3 |
| No. of secondary schools (government and private) | 728 | 2 | 1 | 1 | 2 | 3 | 1 |
| No. of senior secondary schools (government and private) | 728 | 1 | 1 | 0.4 | 1 | 1 | 1 |
| No. of healthcare facilities | 728 | 6 | 5 | 3 | 5 | 8 | 5 |
| **Household characteristics** |  |  |  |  |  |  |  |
| Houses with good living conditions | 736 | 176852 | 202171 | 60274 | 123353 | 214229 | 153955 |
| % houses with roofs made of pucca material | 734 | 82 | 17 | 76 | 87 | 94 | 18 |
| % houses with walls made of pucca material | 734 | 54 | 26 | 32 | 57 | 75 | 42 |
| % houses with floors made of pucca material | 734 | 45 | 27 | 19 | 40 | 69 | 50 |
| % houses owned at the district level | 734 | 88 | 11 | 83 | 91 | 96 | 12 |
| % houses with drinking water from a treated tap water source | 734 | 28 | 22 | 10 | 22 | 41 | 32 |
| % houses that use electricity as source of lighting | 734 | 67 | 28 | 43 | 77 | 91 | 48 |
| % houses with latrine facility on premises- with improved facilities | 734 | 41 | 23 | 22 | 37 | 59 | 37 |
| % houses with bathing facilities on house premises | 734 | 38 | 25 | 16 | 32 | 58 | 42 |
| % houses with bathing facilities connected to a closed drainage | 734 | 13 | 15 | 4 | 8 | 15 | 11 |
| % houses with a kitchen inside the premises | 734 | 58 | 22 | 40 | 59 | 78 | 38 |
| % houses that use LPG as cooking fuel | 734 | 25 | 19 | 9 | 19 | 35 | 25 |
| % houses with any of the specified assets | 734 | 79 | 13 | 71 | 81 | 90 | 18 |
| % houses with banking facilities | 734 | 57 | 17 | 45 | 57 | 69 | 24 |
| **Population characteristics** |  |  |  |  |  |  |  |
| Urban population | 715 | 527370 | 1103236 | 87625 | 248462 | 541660 | 454035 |
| Rural population | 728 | 1144962 | 961395 | 479076 | 940922 | 1501873 | 1022797 |
| % of population between 0-6 years of age | 737 | 14 | 3 | 11 | 14 | 16 | 5 |
| % of scheduled caste or tribe population | 737 | 34 | 24 | 18 | 26 | 40 | 22 |
| % of literate men | 737 | 35 | 5 | 32 | 36 | 39 | 7 |
| % of literate women | 737 | 27 | 6 | 22 | 26 | 31 | 9 |
| % of working men | 737 | 27 | 3 | 26 | 28 | 29 | 3 |
| % of working women | 737 | 14 | 6 | 9 | 14 | 19 | 10 |

\* per 10,000 population in urban areas
\*\* per 10,000 population in rural areas
<sup>1</sup> Sex ratio calculated as number of females per 1000 males

##### NFHS-4

The median percentage (IQR) of households with clean fuel for cooking and an improved sanitation facility was 34% (19-54%) and 47% (31-65%), respectively, an improvement from the Census of 2011 estimates. The median (IQR) percentage of households with an improved drinking water source was 92% (82-97%), and electricity was 95% (85-98%). The average percentage of institutional births was 83% (68-93%), deliveries conducted by health personnel were 85% 972-94%), and births registered for children under 5 years of age were 86% (69-96%).

#### b. Distribution of Health Outcomes in the National Family Health Survey-4 (**Table 2**)

Among children under 5 years, 36% (28-43%) [median (IQR)] had stunting, 33% (23-41%) were underweight, 20% (15-25%) had wasting, and 7% (5-10%) had severe wasting. Among women, on average, 23% (15-29%) had a body mass index below normal (or were underweight), an indication of undernutrition, and 17% (12-25%) were overweight or obese. These numbers suggest a high prevalence of malnutrition among women and children. Among men and women, the median (IQR) prevalence of cardiovascular risk factors was low: women with very high blood sugar 2% (2-3%), men with very high blood sugar 3%(2-5%), women with mildly elevated blood pressure 7% (5-9%), and men with mildly elevated blood pressure 10% (8-15%).

**Table 2:**
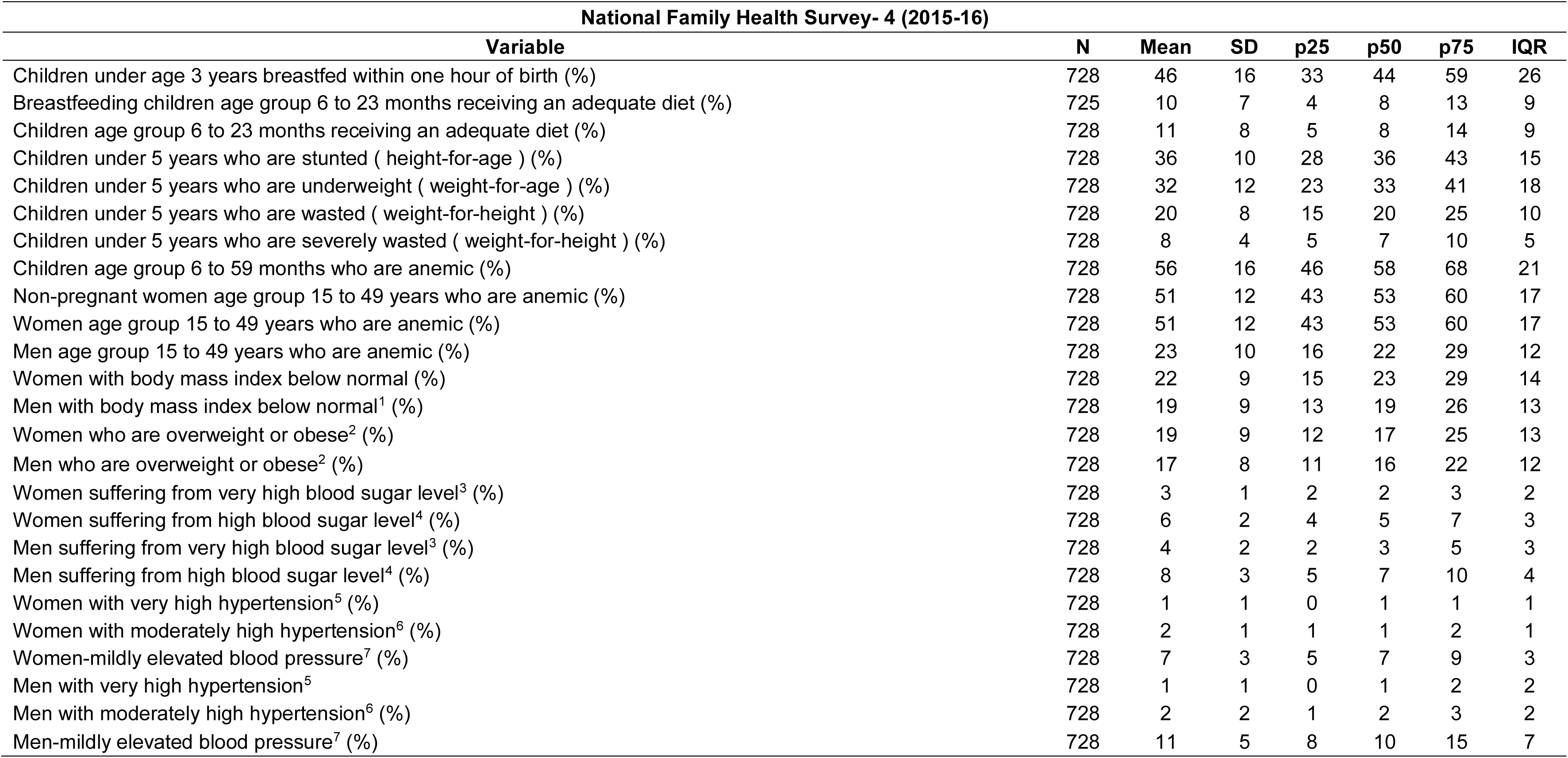
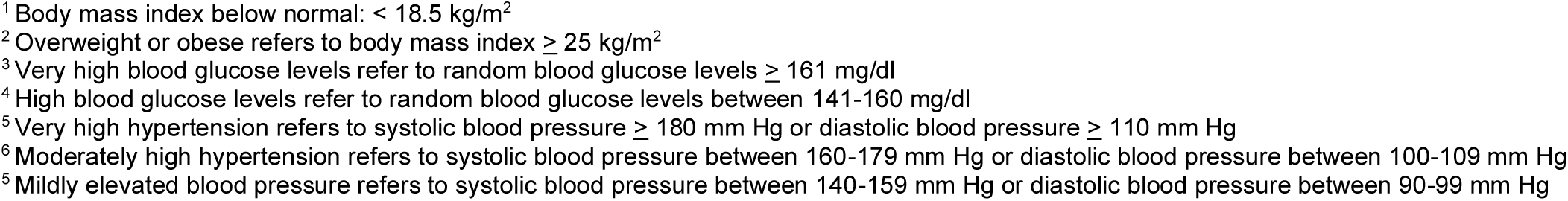
Distribution of health outcomes at the district level in the National Family Health Survey-4.

#### c. Index scores-range, factor loadings

The PCA-generated scores ranged between -4.89 to 7.07 in the census and -8.12 and 5.18 in the NFHS-4. A PCA analysis was conducted in each data source on the standardized indicators listed in **Table 1**. Factor loadings (**Table 3**) indicate the correlation between the indicator and the index score. The magnitude of the factor loading demonstrates the contribution of the variable to the component, and the sign indicates the direction of the relationship between the variable and the component.

**Table 3:**
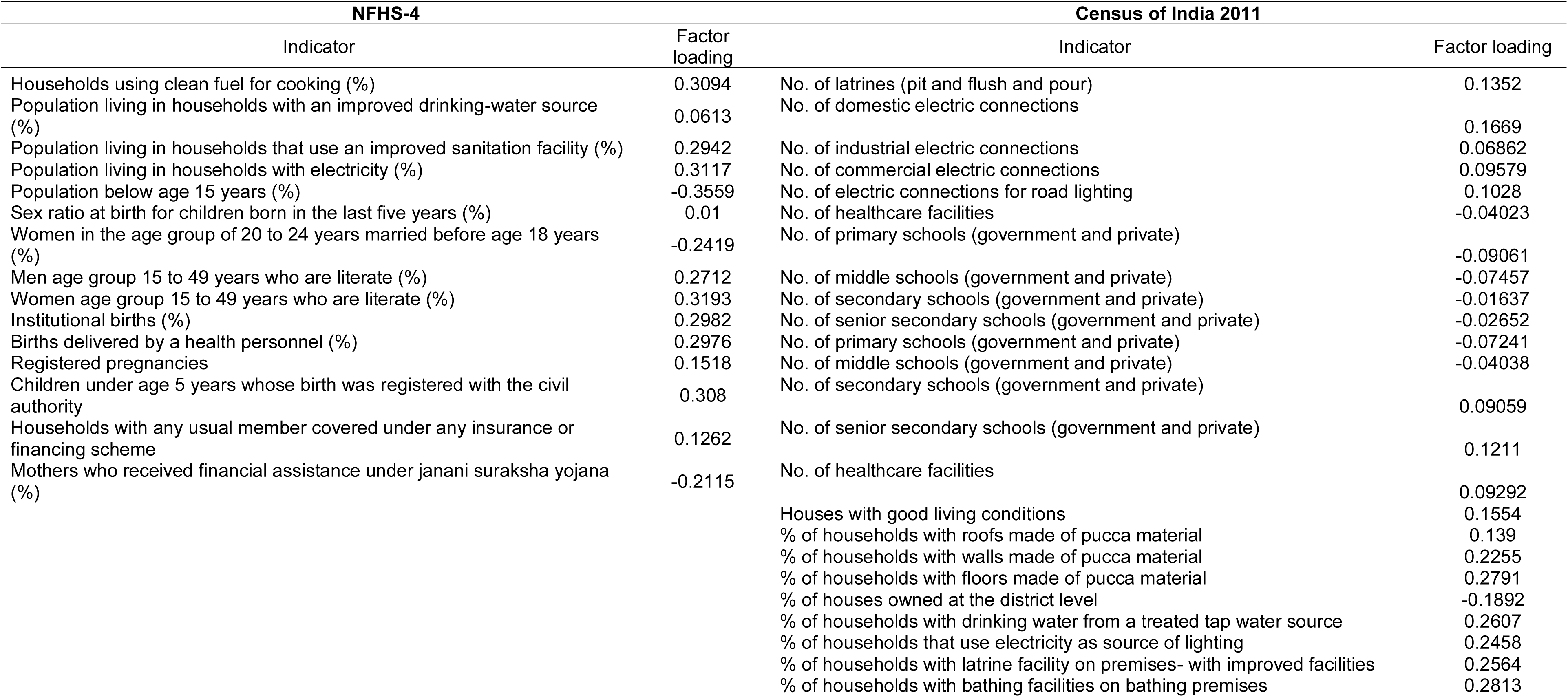
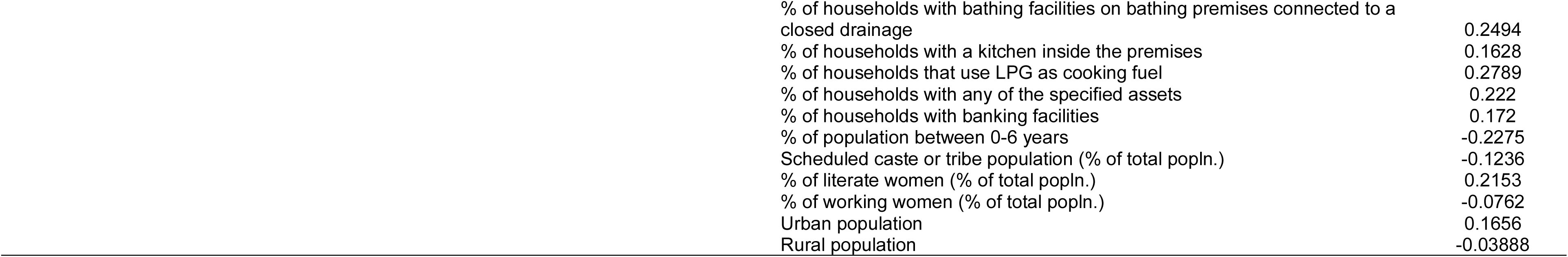
Factor loadings of the first principal component of district-level socioeconomic development scores.

| NFHS-4 |  | Census of India 2011 |  |
| --- | --- | --- | --- |
| Indicator | Factor loading | Indicator | Factor loading |
| Households using clean fuel for cooking (%) | 0.3094 | No. of latrines (pit and flush and pour) | 0.1352 |
| Population living in households with an improved drinking-water source (%) | 0.0613 | No. of domestic electric connections | 0.1669 |
| Population living in households that use an improved sanitation facility (%) | 0.2942 | No. of industrial electric connections | 0.06862 |
| Population living in households with electricity (%) | 0.3117 | No. of commercial electric connections | 0.09579 |
| Population below age 15 years (%) | -0.3559 | No. of electric connections for road lighting | 0.1028 |
| Sex ratio at birth for children born in the last five years (%) | 0.01 | No. of healthcare facilities | -0.04023 |
| Women in the age group of 20 to 24 years married before age 18 years (%) | -0.2419 | No. of primary schools (government and private) | -0.09061 |
| Men age group 15 to 49 years who are literate (%) | 0.2712 | No. of middle schools (government and private) | -0.07457 |
| Women age group 15 to 49 years who are literate (%) | 0.3193 | No. of secondary schools (government and private) | -0.01637 |
| Institutional births (%) | 0.2982 | No. of senior secondary schools (government and private) | -0.02652 |
| Births delivered by a health personnel (%) | 0.2976 | No. of primary schools (government and private) | -0.07241 |
| Registered pregnancies | 0.1518 | No. of middle schools (government and private) | -0.04038 |
| Children under age 5 years whose birth was registered with the civil authority | 0.308 | No. of secondary schools (government and private) | 0.09059 |
| Households with any usual member covered under any insurance or financing scheme | 0.1262 | No. of senior secondary schools (government and private) | 0.1211 |
| Mothers who received financial assistance under janani suraksha yojana (%) | -0.2115 | No. of healthcare facilities | 0.09292 |
|  |  | Houses with good living conditions | 0.1554 |
|  |  | % of households with roofs made of pucca material | 0.139 |
|  |  | % of households with walls made of pucca material | 0.2255 |
|  |  | % of households with floors made of pucca material | 0.2791 |
|  |  | % of houses owned at the district level | -0.1892 |
|  |  | % of households with drinking water from a treated tap water source | 0.2607 |
|  |  | % of households that use electricity as source of lighting | 0.2458 |
|  |  | % of households with latrine facility on premises- with improved facilities | 0.2564 |
|  |  | % of households with bathing facilities on bathing premises | 0.2813 |

|  |  |
| --- | --- |
| % of households with bathing facilities on bathing premises connected to a closed drainage | 0.2494 |
| % of households with a kitchen inside the premises | 0.1628 |
| % of households that use LPG as cooking fuel | 0.2789 |
| % of households with any of the specified assets | 0.222 |
| % of households with banking facilities | 0.172 |
| % of population between 0-6 years | -0.2275 |
| Scheduled caste or tribe population (% of total popln.) | -0.1236 |
| % of literate women (% of total popln.) | 0.2153 |
| % of working women (% of total popln.) | -0.0762 |
| Urban population | 0.1656 |
| Rural population | -0.03888 |

In NFHS-4, the first component explained 42% of the total variance; loadings ranged from -0.36 to 0.32. Indicators that contributed to the final score at factor loading > 0.20 included the percentage of households with clean fuel for cooking; improved sanitation; and electricity; the percentage of the population less than 15 years (negatively correlated); women aged 20-24 years who wed before 18 years (negatively correlated); percentage of literate men and women; institutional births and deliveries conducted by health personnel; registered births and mothers who received financial assistance through Janani Suraksha Yojana (negatively correlated). On excluding weakly correlated indicators, the first component explained 55% of the variance, and the Kaiser-Meyer-Olkin (KMO) statistic indicated adequate sampling.

In Census data, the first component explained 29% of the total variance, and factor loadings ranged between -0.23 and 0.28. Indicators that were correlated with the final index score (factor loading > 0.20) were the percentage of households with pucca walls or floors; an improved drinking water source; improved sanitation; electricity; bathroom on premises; closed drainage; LPG for cooking fuel; specified assets; the percentage of children between 0-6 years (negatively correlated) and percentage of literate women. On excluding non-contributing indicators, the first component explained 65% of the variance and the KMO statistic indicated adequate sampling. A summary comparison of factor loadings across each dataset is presented in **Supplemental Table 5.**

### II. Index validation

#### a. Construct validity

##### NFHS-4

Correlations between indicators ranged from -0.78 to 0.96 (**Table 4**). Socioeconomic indicators, such as literacy in men and women, showed strong positive correlations (r=0.70, r=0.78). Among housing characteristics, moderately strong (r= 0.50 to 0.69) positive correlations were observed between the availability of clean cooking fuel, electricity for lighting, and improved sanitation facilities. Improved drinking water was weakly correlated with other housing indicators, suggesting it was a poor housing characteristic indicator. Among community infrastructure indicators, institutional births and births by health personnel had the strongest correlation (r=0.96), with registered births and pregnancies also moderately positively correlated. Financial assistance under the Janani Suraksha Yojana and insurance coverage were the least contributing indicators. Sociodemographic indicators were weakly correlated with each other.

**Table 4:**
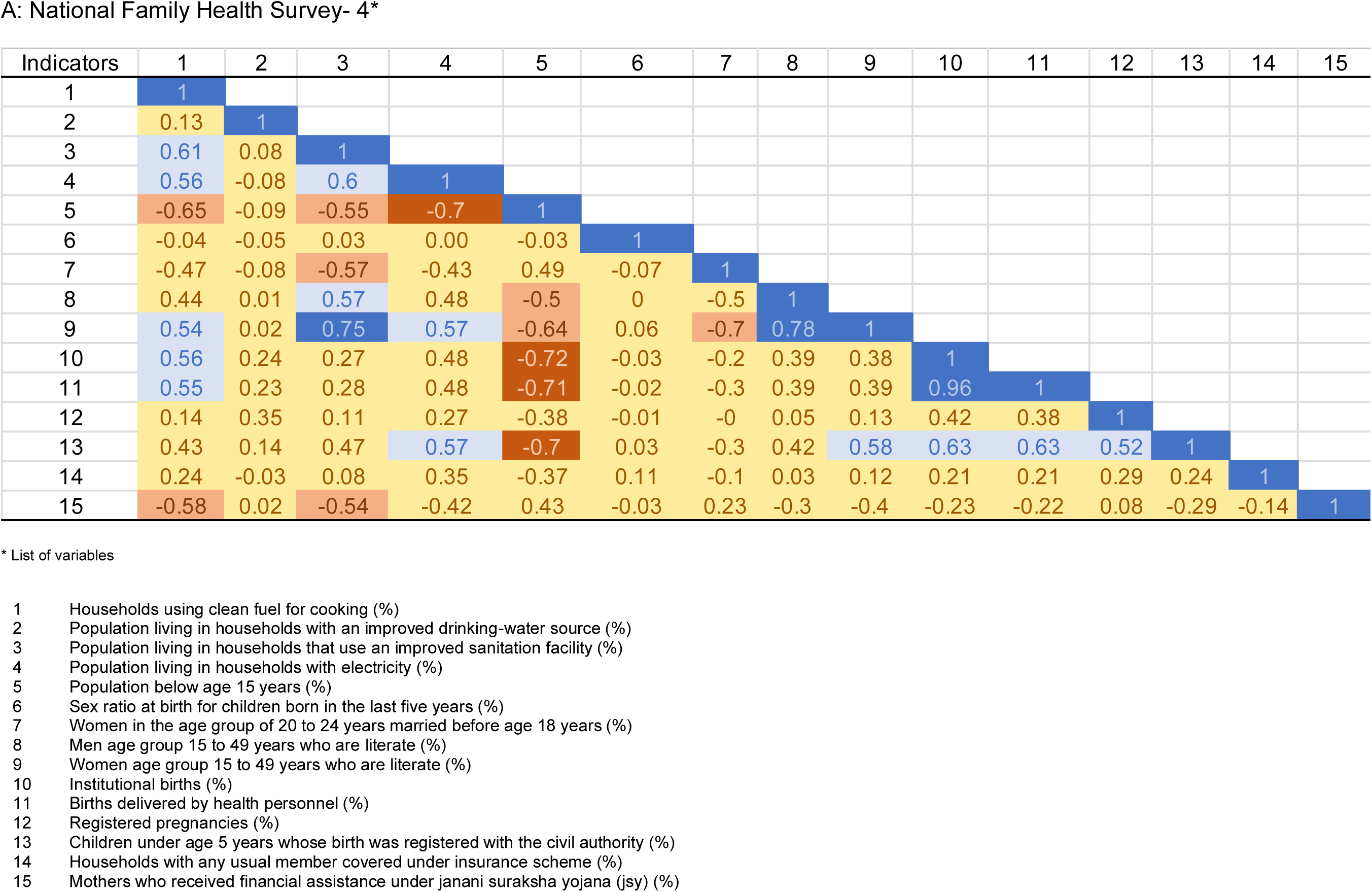
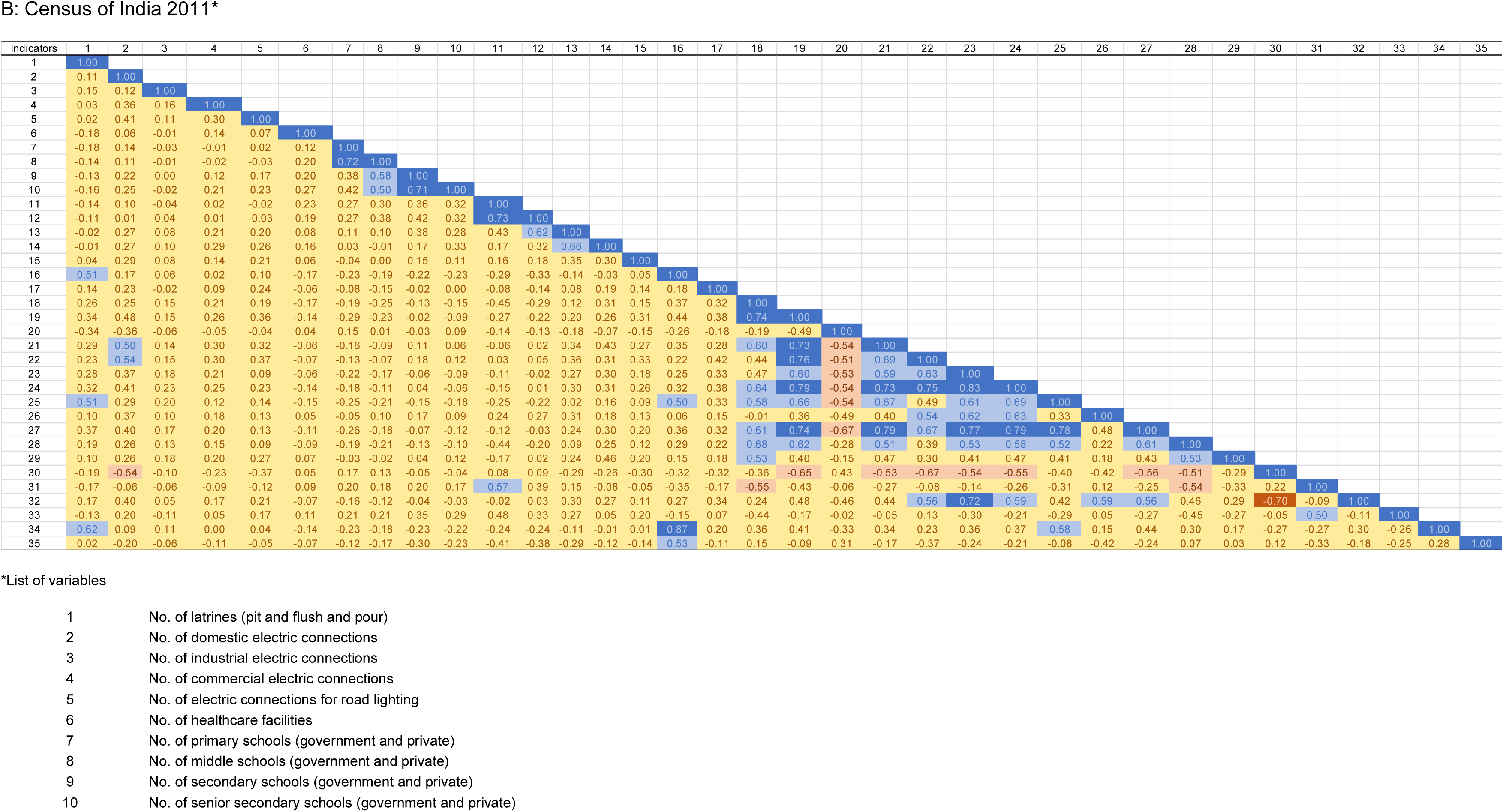
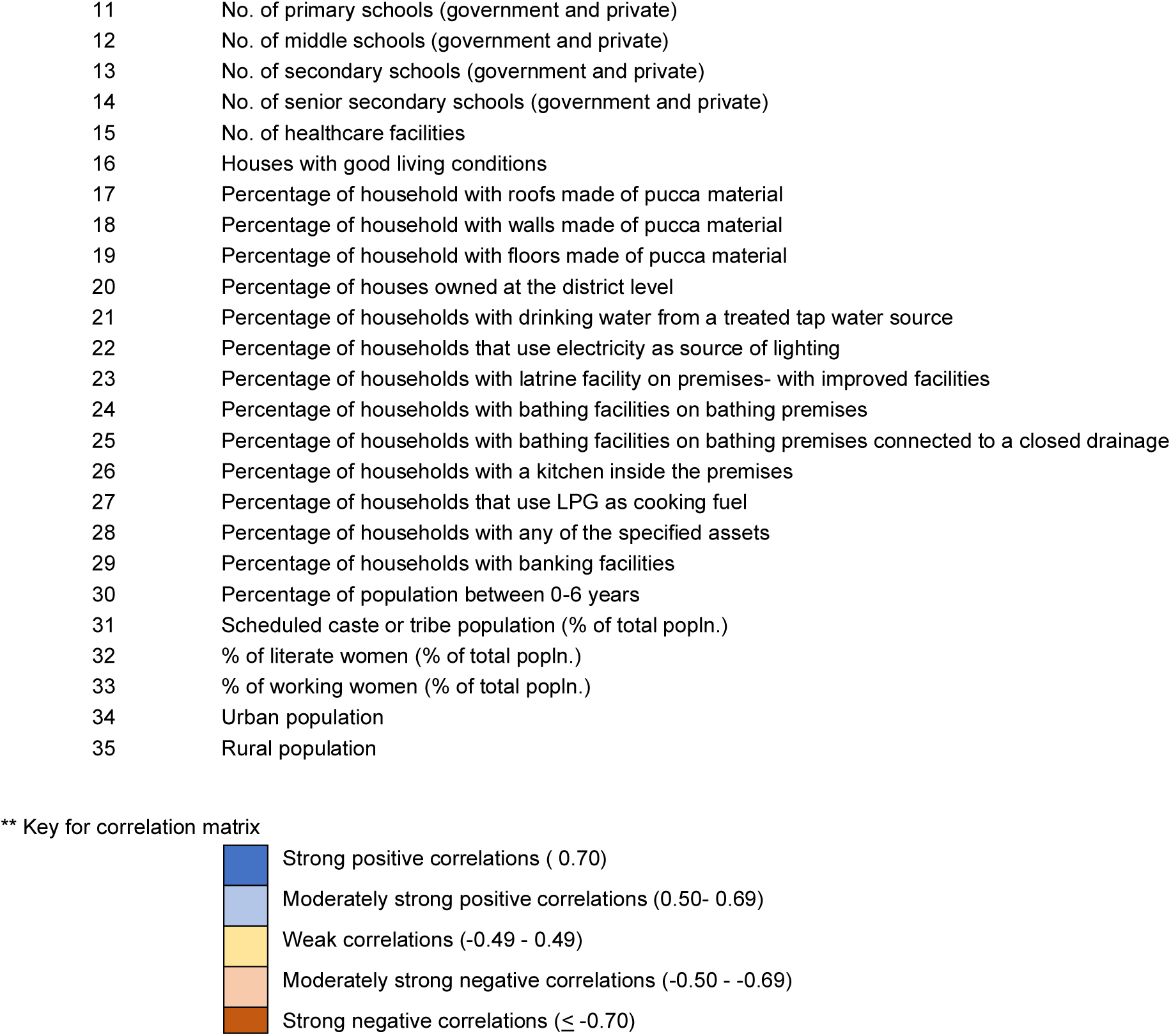
Correlations between indicators in each of the following datasets: A: NFHS-4, B: Census of India 2011**.

##### Census

Correlations between indicators in the census ranged from -0.67 to 0.96 (**Table 4**). Socioeconomic indicators were weakly negatively correlated. Housing characteristics were moderately positively correlated with each other, except for home ownership and access to banking facilities, which were weakly negatively correlated with good housing conditions.

Community infrastructure indicators, such as the number of primary, middle, and high schools, were positively correlated, while other infrastructure indicators were weakly correlated and less contributive. Similar to NFHS-4, sociodemographic indicators were weakly correlated. A higher percentage of children under 6 years was correlated with worse housing characteristics.

#### b. Criterion validity

##### Concurrent criterion validity-Pearson correlations between scores within the same dataset

The PCA and percentile-generated scores were strongly positively correlated within both the NFHS-4 (r=0.86) and the Census (r=0.81) data. Within each dataset (**Table 5**), when evaluating domain-specific scores, percentile and PCA-derived domain scores in the NFHS were strongly positively correlated. In the Census, all other domain scores were poorly correlated except for the percentile and PCA domain scores for housing indicators, which were strongly positively correlated.

**Table 5:**
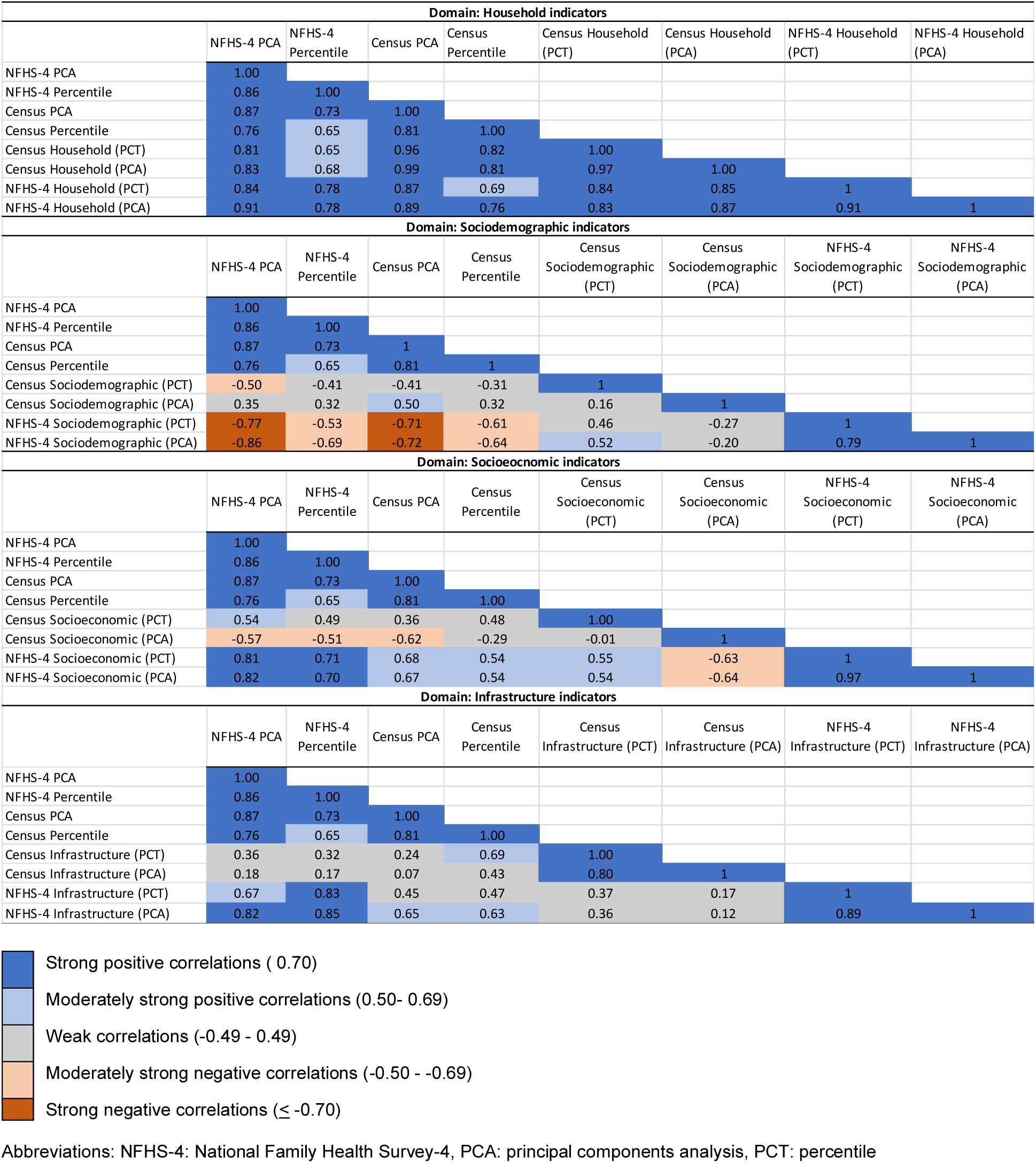
Correlations between domains of district-level socioeconomic scores across the Census and the National Family Health Survey-4.

##### Concurrent criterion validity-Pearson correlations between domains across datasets

Household indicators were moderately-strongly positively correlated between the NFHS-4 and Census data, using either a PCA or percentile scoring system (**Table 5**). For sociodemographic indicators, the NFHS-4 percentile-derived score was moderately positively correlated (r=0.52) with the Census percentile score. There were less consistent correlations in this domain. Among socioeconomic indicators, the census percentile domain score was moderately positively correlated with the NFHS-4 domain scores. Conversely, the census PCA-derived domain score was moderately negatively correlated with the NFHS domain scores. The difference in the Census domain scores was the inclusion of the percentage of working women in the percentile score. The infrastructure indicators were not correlated across datasets.

##### Comparison of district-level socioeconomic development maps by index source data and methodology

All four indices generated in this analysis (**Figure 2**) suggest that the lowest socioeconomic development was largely in the central part of the country, extending from Rajasthan in the northwest to Manipur, Nagaland, and parts of Arunachal Pradesh in the East. Areas with the highest socioeconomic development were concentrated in the south, in Maharashtra, Karnataka, Kerala, Tamil Nadu, and parts of Andhra Pradesh. Within data source, the NFHS-4 percentile index classified more districts in Jammu and Kashmir, western parts of Andhra Pradesh, and eastern parts of Karnataka as less socioeconomically developed (compared to the NFHS-4 PCA score). The PCA-generated census score identified noticeably more widespread areas of low socioeconomic development in the central, northeast, and Jammu and Kashmir than the census percentile rank score.

**Figure 2:**
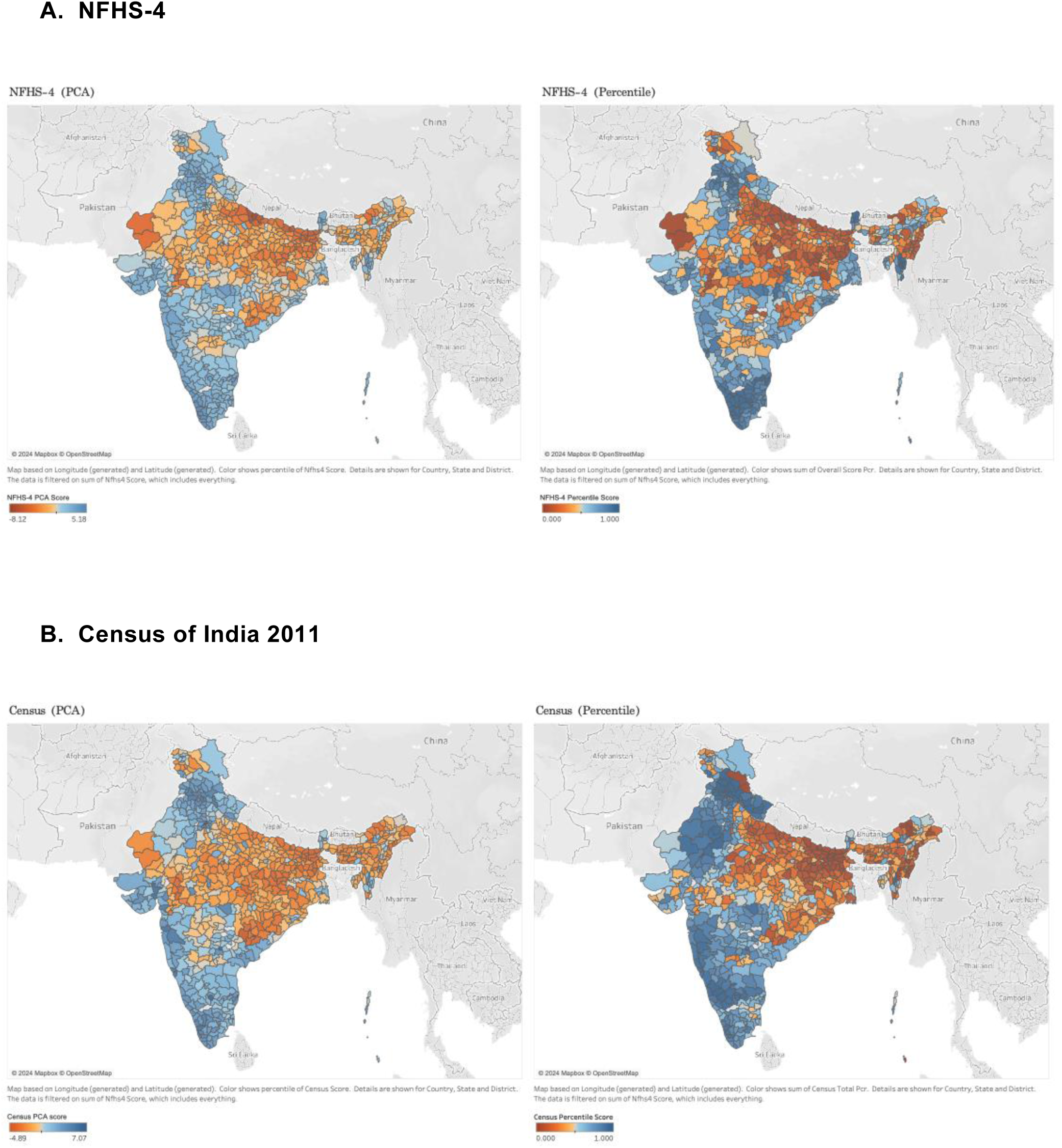
District-level socioeconomic development scores by data source and method

##### Comparison of district-level socioeconomic development maps with the Multidimensional Poverty Index

Comparing maps of the four scores (**Figure 2**) with the Multidimensional Poverty Index^61^, we observed all scores were closely aligned in identifying districts with lower socioeconomic development in the central belt and parts of Andhra Pradesh. Unlike in the Multidimensional Poverty Index, the NFHS-4 derived scores and census PCA score identified areas of low socioeconomic development in Jammu and Kashmir while the percentile census score showed several districts in the East as having low socioeconomic development. Like the Multidimensional Poverty Index, the census percentile score identified large parts of Jammu and Kashmir as being relatively socioeconomically advantaged.

##### Predictive validity

Bivariate Pearson correlations were explored between health outcomes in the NFHS-4 and each of the four overall district-level socioeconomic development scores (**Table 6**). The percentage of children under 5 years who were stunted (height-for-age) was negatively correlated with all four scores. The percentage of children under 5 years who were underweight (weight-for-age) was moderately negatively correlated with the NFHS-4 and Census PCA score (r=-0.57, r=-0.50). The percentage of women who were underweight was moderately negatively correlated with the PCA and percentile NFHS-4 scores (r=-0.57, r=-0.55) and the Census PCA score (r=-0.50). This suggested that undernutrition was associated with lower socioeconomic development. The percentage of women and men who were overweight or obese was moderately-strongly positively associated with all four scores. All the other evaluated health outcomes were either not correlated or weakly correlated with the socioeconomic development indices. The correlations between urban and rural-specific scores with health outcomes were limited and described in the supplement (**Supplemental Tables 6, 7**).

**Table 6:**
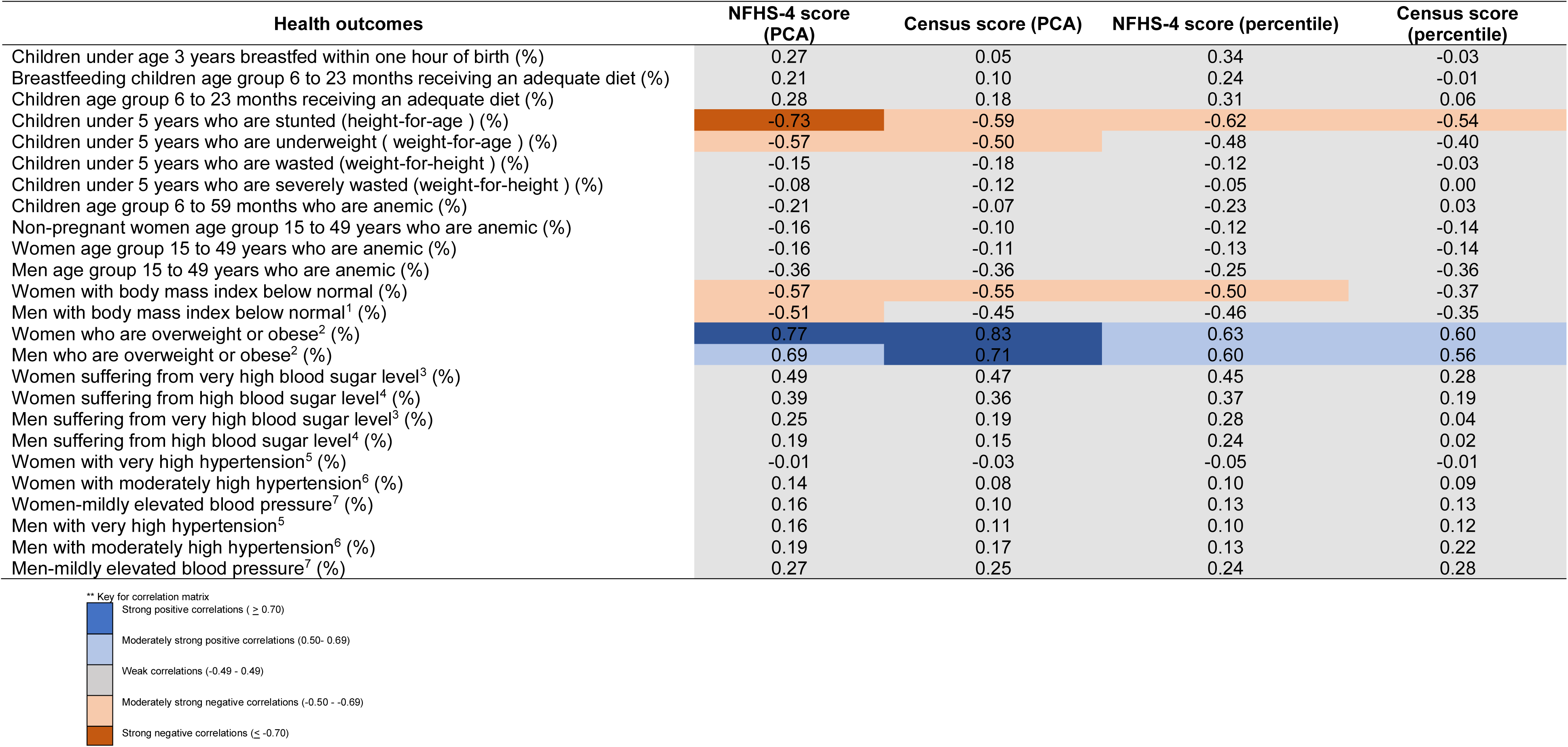
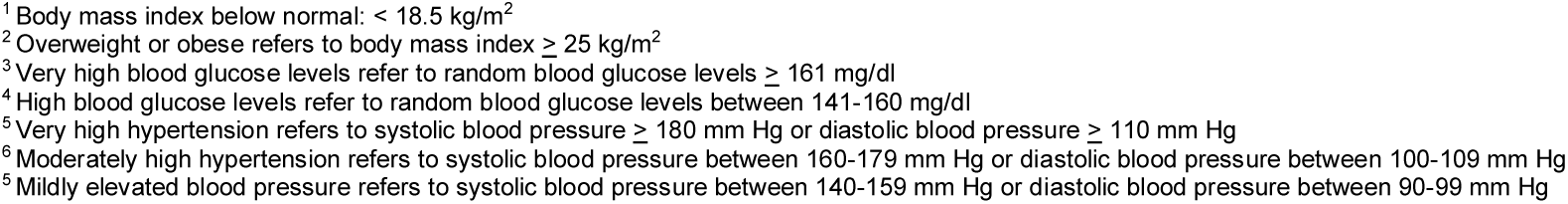
Correlations between district-level socioeconomic development scores and health outcomes in the National Family Health Survey-4*\**.

## Discussion

In this study, we developed district-level indices for socioeconomic development in India and evaluated their construct and criterion validity. Using two different methodologies applied to two separate publicly available data sources -- principal components analysis (PCA) and percentile ranking -- we generated four distinct indices. While both PCA and percentile ranking methods within the NFHS-4 data yielded similar results, PCA was limited by component interpretability, sensitivity to outliers, and subjective component selection. In contrast, percentile ranking offered a reproducible, straightforward approach, using intentionally selected indicators based on their relationship to socioeconomic development and health outcomes. All four indices were largely consistent in classification of areas as socioeconomically developed or underdeveloped. The NFHS-4 dataset proved particularly robust, with strong construct validity confirming the chosen variables’ ability to capture socioeconomic development. Scores from NFHS-4 exhibited strong internal and external concurrent validity compared with the multidimensional poverty index.

These scores effectively predicted key nutritional health outcomes among children and adults. The NFHS-4 percentile score provided a comprehensive assessment of overall district-level socioeconomic development. However, neither dataset nor method demonstrated good predictive validity when stratifying by urban and rural areas, indicating the need for further refinement to develop urban-rural specific scores.

The domains selected for this analysis—socioeconomic, sociodemographic, housing, and infrastructure characteristics—align with previous indices and have been associated with health outcomes or determinants of health.^18,19,24,62,63^ Socioeconomic indicators like education and employment reflect social standing and influence health through pathways such as literacy and income.^64–67^ Housing and infrastructure, including quality of living and access to public amenities, are critical indicators of socioeconomic development and environmental vulnerability.^33,35,68^ Prior studies often overlooked informal, unpaid, or low-paid work, especially among women.^67^ This analysis includes socioeconomic indicators for both men and women to address some of these gaps. It also included information on sociodemographic indicators, such as the proportion of vulnerable groups like elderly people, gender-disaggregated indicators, and religious and/or ethnic minorities, that face structural barriers that impact their access to resources and health. Among demographic characteristics, our study found that a higher percentage of a younger population at the district-level was generally associated with lower community-level socioeconomic development across both the data sources. We did not include any health outcomes as indicators, which prior studies have done.^24,62,69^ We hypothesized strong positive correlations among related indicators in the same domain. The NFHS-4-derived scores demonstrated good construct validity, confirming the selected domains and indicators effectively measure socioeconomic development.

The absence of a gold standard method for creating indices of socioeconomic development has led to the use of various methodologies. The Multidimensional Poverty Index (MPI), adopted by the Government of India, uses the Alkire-Foster methodology to measure socioeconomic deprivation across health, education, and standard of living.^22^ It includes 12 indicators with differential weights across these three equally weighted dimensions. The MPI is calculated as the product of the proportion of multidimensionally poor people and the intensity of poverty. The MPI offers strengths such as multidimensionality and insights into both the incidence and intensity of poverty.

Like the percentile ranking method, the MPI assigns ranks based on deprivation across multiple indicators as does the percentile ranking method. The MPI and PCA both assign weights to indicators. The PCA determines weights statistically based on variance and the MPI assigns fixed weights. The MPI with its fixed dimensions can overlook other contextually relevant domains, and cut-off values often lack a strong empirical basis, potentially impacting precision and policy decisions. The discretization of indicators and their distribution can also meaningfully impact the MPI and poverty assessments.^70^ The MPI does not account for deprivation faced by specific vulnerable groups, such as ethnic minorities or persons with disabilities, providing an incomplete assessment of socioeconomic deprivation.

Principal components analysis is the most commonly applied method to develop area-level scores for socioeconomic composition. PCA reduces selected indicators to uncorrelated components that explain variance in the dataset.^71–73^ Due to its assumptions of linearity and equal intervals between variables, it is ideal for continuous data, and this poses a challenge for using categorical variables commonly found in national surveys like the ones used in the present analysis. Additionally, scores are developed from the data-driven weight of variables and not based on their contribution to socioeconomic development. In the present analysis, we have tried to overcome this feature by using variables that have previously been demonstrated to represent socioeconomic development and were associated with health outcomes. The percentile ranking method assigns equal weights to indicators, potentially misrepresenting those with greater socioeconomic impact.^33^ Despite this, it is simple to compute and accessible, ensuring comparability across indicators by considering the relative position within its distribution. The resulting index is customizable and interpretable, offering clear insights into relative deprivation levels across different areas or population groups.

Validation is crucial for assessing the relevance of constructed indices, yet it is often overlooked due to the absence of a gold standard method. The methods used are correlations and chi-square tests against existing indices (e.g., DHS wealth index), Kappa statistic, and Bland Altman curves.^74–77^ In the present study, all four scores demonstrated good internal and external concurrent criterion validity to the extent it could be evaluated. Areas of low socioeconomic development identified in this study concurred with prior work, including the Multidimensional Poverty Index, conducted at the state or district level and consistently identified regions in the central and northeastern parts of the country as areas with the lowest development.^19,20,78–81^ In contrast, areas to the north and south had higher development.^19,20,78–81^

Our scores also demonstrated good predictive validity in identifying health inequalities. Prior work focusing on health outcomes such as healthcare utilization, childhood morbidity, mortality, nutrition, and COVID-19 have demonstrated an association with area-level socioeconomic development; however, few were validated.^23,24,63,77,82–87^ In the present study, in areas with higher socioeconomic development, there was a greater prevalence of overweight and obesity among adults, elevated blood pressure in women, and a lower prevalence of undernutrition in children. These findings reflect the ongoing health transition in a country like India, where metabolic disorders are seen in groups with greater socioeconomic gains. Unlike the overall score, the urban-rural stratified scores did not adequately predict health outcomes. This needs further investigation, though the NFHS-4-derived scores demonstrated good construct and concurrent validity.

This study had several strengths and limitations that warrant consideration. Its strengths include a multidimensional approach to evaluating socioeconomic development across four well-established domains, utilizing diverse data sources and two analytical approaches. However, the data used were nearly a decade old, potentially not representing current socioeconomic development. While the census data provided extensive information on infrastructure, only data on educational and healthcare facilities were analyzed due to missing information. The study included age and gender-disaggregated indicators, but the absence of healthcare quality and health literacy indicators was a limitation. The analysis could be susceptible to the Modifiable Areal Unit Problem (MUAP), which can obscure localized spatial patterns; however, access to smaller geographic units was unavailable in the NDAP to test for this issue.^88,89^ Despite this, our study stands out as one of the few in India that evaluated the construct and predictive validity of the indices. However, the absence of an expert panel or Delphi method in the validation process limited context-specific insights. Despite this, the study used variables observed to capture different aspects of socioeconomic development in previous studies and demonstrated good construct validity in at least one dataset. Lastly, the inability to recreate the Multidimensional Poverty Index using available NDAP data precluded the evaluation of concurrent and predictive validity by examining correlations.

## Implications and Conclusion

Developing socioeconomic development indices involves challenges such as data scarcity and quality, selecting and weighting indicators, methodological decisions for normalization and aggregation, capturing temporal and spatial variations, addressing cultural sensitivity, and ensuring equity and inclusivity. The dynamic nature of socioeconomic development and the absence of a standardized approach further compound these challenges. In this analysis, indices were constructed and validated using well-accepted methods. Data from the NFHS-4 demonstrated good construct validity, supporting the chosen dimensions and the utility of NFHS data. While the census provides comprehensive information, selecting appropriate socioeconomic indicators may require combining theory-based and data-driven approaches, especially for urban-rural stratified scores.

The percentile and principal components analysis methods in the NFHS-4 data produced similar results. Percentile ranking offers a reproducible and straightforward approach, using carefully selected indicators related to socioeconomic development and health outcomes. The NFHS-4 percentile score demonstrated good concurrent validity compared to the NFHS-4 PCA score.

However, scores from both methods had similar relationships with health outcomes, with the PCA scores having stronger correlations. Future work should incorporate other indicators with a demonstrated relationship with health behaviors and outcomes, such as the availability of green spaces, air and water quality indicators, and social cohesion. Incorporating some of these indicators might improve the prediction of health outcomes by these indices. A potential approach to indicator selection is to first identify the most variable indicators using PCA and then apply percentile ranking to create an index score. This method leverages the strengths of both techniques, ensuring a more robust selection process. The persistence of low socioeconomic development in the central belt of India highlights the need for a deeper investigation using both qualitative and quantitative methods.

The indices in this analysis allow for investigating the intersectionality of various dimensions of socioeconomic development, considering how multiple factors (e.g., gender, ethnicity, age) may interact to shape an individual’s experiences. The relationship between socioeconomic development and health outcomes in this study does not establish causality and needs to be explored in future temporal and spatial analyses. Further work will also be needed to assess the utility of indices of socioeconomic development in the clinical setting.

## Data Availability

Datasets used in this analysis, including dataset number and link, have been provided in Supplemental Table 1.

