## Supplement for "Development and validation of a district-level index of socioeconomic development for India"

**Supplemental Methods**

Data Source: National Data Analytics Platform

The National Data Analytics Platform (NDAP), an initiative spearheaded by NITI Aayog, an Indian government-run public policy think tank, facilitates public access to government survey data conducted by various ministries, employing diverse methods at different time points, all of which were standardized.[1] Merging datasets in the NDAP was done using common dimensions, either geographic (state, district, and sub-district) or temporal units (year and month). Using local government directory codes[2] corresponding to administrative units aligned with current Indian geography, datasets from previous periods were transformed to match the latest geographic units through weighted merging or splitting of previous units, as appropriate.[1] Further details on the standardization process have been published previously.[1]

Indicators

**Sociodemographic indicators** included population below 15 years, sex ratio at birth, and early marriage rates from NFHS-4, and population below 6 years, urban/rural populations, and scheduled caste/ tribe data from the Census. These indicators indicate vulnerabilities to ill health due to age-related illnesses, lack of financial security, and/or other systemic barriers.[3,4] **Socioeconomic indicators** included literacy rates and employment classification from the Census and literacy rates from NFHS-4, as education influences health through income, knowledge, and decision-making pathways.[5-7] **Housing indicators in the Census and NFHS-4** covered household standard of living characteristics and assets, which are indicators of wealth and generational wealth.[8] **Community infrastructure and resource indicators** included the number of schools from the Census and healthcare facilities and civil registration services from NFHS-4. Community-level exposures, whether environmental (e.g., air or water pollution), physical (e.g., availability of public transportation, neighborhood green spaces), or social (e.g., crime and safety) can have an independent effect on health.[9-12]

**Health outcomes** from NFHS-4 included nutritional indicators and cardiovascular risk factors. Nutritional indicators included infant breastfeeding, childhood wasting, stunting, and anemia, as studies have shown an inverse association between these indicators and socioeconomic development.[13-19] Cardiovascular risk factors included hypertension, diabetes mellitus and being overweight or obese. Socioeconomic disadvantage is associated with a higher prevalence of behavioral risk factors (e.g., unhealthy diet, tobacco smoke). In comparison, socioeconomic advantage is associated with a higher prevalence of obesity, hypertension, diabetes, dyslipidemia, and physical inactivity.[20-27]

Unit of analysis

India presently has 28 states and 8 union territories, that are further subdivided into smaller administrative units for ease of governance. These administrative units include divisions (clusters of districts), districts, sub-districts, and more. The district, headed by a district magistrate, usually a civil service appointee, is an administrative sub-division. The formation of districts is not based on population size and is at the discretion of the state governments. In this study, the district was chosen as the unit of analysis due to the availability of several datasets at this level of granularity. The NDAP data was standardized to provide information on up to 765 districts, according to present geography.

Analytic methods for urban-rural scores

As in the primary analysis, two methods were used to construct scores for district-level urban and rural areas- principal component analysis and percentile ranking. For the NFHS-generated scores, the same indicators as in the primary analysis were used and stratified by urban-rural area. For the census-derived scores, urban areas did not contain indicators from villages (representative of rural areas), and similarly, rural areas did not contain indicators from towns (representative of urban areas). All other indicators were the same. In the NFHS-derived scores, 15 indicators were included; in the Census-derived urban score, 30 indicators were included; and in the census-derived rural score, 25 indicators were included. Validation methods for the stratified scores included construct and predictive validity. Concurrent validity could not be assessed due to the unavailability of district-level urban-rural stratified maps of the Multidimensional Poverty Index (MPI).

**Supplemental Results**

I. Distribution of indicators at the district level in urban and rural areas- Across both data sources, in urban areas, more homes had better housing conditions. In rural areas, the sex ratio was higher (922 females per 1000 males), a greater number of schools and higher home ownership was observed (**Supplemental Table 3**)

a. Census: Rural areas in districts had a greater number of primary and middle schools as compared to urban areas. A greater number of homes in urban areas were built of pucca materials and had better facilities, while there was greater home ownership in rural areas. There was a higher percentage of working women in rural areas and literate women in urban areas.

B: NFHS-4: In the NFHS-4, for urban areas, we restricted the districts to those with an urban population of at least 50%. Urban areas had more homes with a clean source of cooking fuel and an improved sanitation facility. Rural areas (922 females per 1000 males) had a higher sex ratio than urban areas and more women who were wed before 18 years of age. Urban areas had a greater number of births conducted by trained personnel, institutional births, and registered births. More urban homes had a member who had health insurance while rural areas had higher utilization of government schemes for safe motherhood.

I. Index Construction

a. Urban-Rural Index scores- range, factor loadings

In rural areas, the first component of the PCA analysis explained 51% of the variance in the NFHS-4 data. The factor loadings of the PCA-based scores using NFHS-4 data were all positive and ranged from 0.05 to 0.33. The indicators that were positively correlated ($\geq$0.20) with the final score were the percentage of; literate men, literate women, institutional births, deliveries conducted by health personnel, registered pregnancies, registered births, households with an improved drinking water source, households with electricity, an improved sanitation facility and, sex ratio at birth. The indicators that contributed the least were the percentage of; households with clean fuel for cooking, the population below 15 years of age, women aged 20-24 years who wed before 18 years of age, household members covered by an insurance scheme and registration of mothers in the Janani Suraksha Yojana scheme. On excluding the indicators that contributed least to the final score, the first component explained 69.7% of the variance. The KMO statistic for all the retained variables was over 0.80, indicating good sampling adequacy.

Using census data, restricted to rural areas, the first component of the PCA explained 31.5% of the variance. The factor loadings ranged from -0.25 to 0.31. Indicators that contributed the most (factor loading $\geq$ 0.20) to the final score included the percentage of; households with pucca walls and floors, households with an improved water source, households with electricity, households with improved sanitation, households with bathrooms on the premises, households with closed drainage, households with LPG gas connection, households with the specified list of assets, households with members who have bank accounts, women who were literate and children aged 0-6 years (negatively correlated). After excluding the non-contributing indicators, the first component explained 53.5% of the variance. Aside from the percentage of households with improved sanitation, all other indicators had a KMO statistic of $\geq$ 0.80.

In the NFHS-derived urban PCA score, the first component explained about 91% of the variance. Factor loadings ranged from 0.21-0.27. The KMO statistic indicated high sampling adequacy (> 0.90); all indicators were retained for the final score. Using the census data, the PCA generated a first component that explained 27.6% of the variance. Factor loadings ranged from -0.23 to 0.31. Indicators that were strongly correlated (factor loading $\geq$ 0.20) to the final score included the percentage of; towns with domestic electric connections, households with pucca floors, households with; an improved drinking water source, improved sanitation, electric connections, bathrooms on premises, kitchens inside the home, closed drainage, LPG as cooking fuel, specified assets and, children in the age group of 0-6 years. On excluding indicators with low factor loadings, the first component of the PCA explained 56.8% of the variance. Factor loadings were $\geq$ 0,20, and KMO statistic $\geq$ 0.80.

### II. Index validation

a. Construct validity

For the NFHS-derived rural score, correlations ranged between -0.38 to 0.98. Among sociodemographics, the sex ratio at birth was moderately positively correlated with the percentage of the population below 15 years of age (r=0.63). The percentage of literate men and women was strongly positively correlated (r=0.86). Indicators in the domain of household characteristics were poorly correlated with each other. Among community infrastructure indicators, institutional births were strongly positively correlated with deliveries conducted by healthcare personnel, registered pregnancies, and registered births. Similarly, registered pregnancies and births and deliveries by healthcare personnel were strongly positively correlated ($r \geq 0.70$). For the Census-derived rural score, correlations ranged between -0.45 to 0.78. Among community infrastructure indicators, the percentage of schools in villages at the district level was strongly correlated. Each level of schooling was strongly positively correlated with the subsequent level of schooling. Among household characteristics, having tap water as a source of drinking water was strongly positively correlated (r=0.70) with having LPG as a source of fuel. Improved sanitation and a bathroom on the premises were also positively correlated (r=0.77). There were no other strong correlations within or between domains.

For urban areas, the NFHS-derived indicators were generally strongly ($r \geq 0.80$) positively correlated with each other within and across the domains with a range of 0.58 to 0.99. The indicators with the weaker correlations were household members covered by health insurance and women who received assistance through the Janani Suraksha Yojana scheme. In contrast to the NFHS-derived indicators, the census indicators were largely poorly correlated, with correlations ranging from -0.61 to 0.97. Community infrastructure indicators were poorly correlated with each other. Among household characteristics, having an LPG connection for cooking fuel was strongly positively correlated with having a bathroom on the home premises.

b. Criterion validity

i. Concurrent validity

The NFHS-4-derived rural scores (PCA and percentile derived) were strongly and positively correlated (r=0.95) with each other, as were the Census-derived rural scores (r=0.89). The NFHS-4-derived PCA and percentile score for urban areas were perfectly correlated (r=1.00). The census derived PCA and percentile urban scores were strongly positively correlated (r=0.93). We could not compare the urban and rural scores to the Multidimensional Poverty Index due to the unavailability of district-level Multidimensional Poverty Index scores that were stratified by urban-rural areas.

ii. Predictive validity

Neither of the NFHS-derived rural scores had strong positive or negative correlations with any of the health outcomes. Both the census rural scores were moderately negatively correlated with childhood stunting and underweight and positively correlated with overweight or obesity in men and women. All four scores correlated poorly with all selected health outcomes in urban areas. When considering the percentage of population in urban areas in a district, 38% of districts had an urban population of ≥25%, 12% of districts had an urban population of ≥50% and 4% of districts had an urban population of ≥ 75%. When restricted to districts with an urban population of 50% or greater, the NFHS-derived scores were moderate- strongly negatively correlated with hypertension in women.

**Supplemental Figure 1**: Domains and indicators for socioeconomic development in the Census of India 2011 and the National Family Health Survey-4

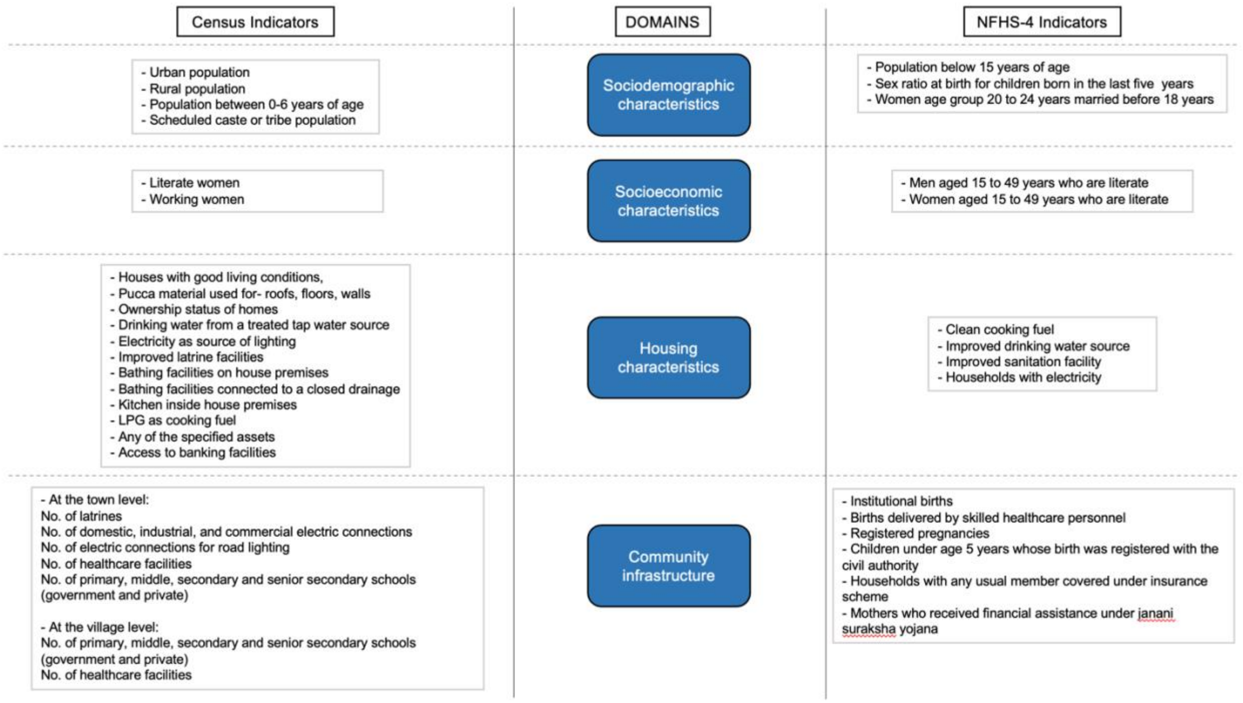

**Supplemental Table 1:** Datasets from the National Data Analytics Platform used in this analysis

| Dataset Name | Dataset Number | Dataset Link |
|---|---|---|
| Primary Population Census 2011 | 6000 | https://ndap.niti.gov.in/dataset/6000 |
| Town Amenities or Directory from Census 2011 | 7135 | https://ndap.niti.gov.in/dataset/7135?tab=profile |
| Village amenities or directory from census 2011 | 7121 | https://ndap.niti.gov.in/dataset/7121 |
| Census 2011: Houses, Household Amenities and Assets Census Houses and the uses to Which they are Put [H-1] | 6993 | https://ndap.niti.gov.in/dataset/6993 |
| Census 2011: Houses, Household Amenities and Assets Households by Availability of Bathing Facility and type of Drainage Connectivity For Waste Water Outlet [HH-9] | 7012 | https://ndap.niti.gov.in/dataset/7012 |
| Census 2011: Houses, Household Amenities and Assets Households by Availability of Separate Kitchen and type of Fuel used For Cooking [HH-10] | 7000 | https://ndap.niti.gov.in/dataset/7000 |
| Census 2011: Houses, Household Amenities and Assets Households by Availability of type of Latrine Facility [HH-8] | 7011 | https://ndap.niti.gov.in/dataset/7011 |

| | | |
|---|---|---|
| Census 2011: Houses, Household Amenities and Assets Households by Main Source of Drinking Water and Location [HH-6] | 7009 | https://ndap.niti.gov.in/dataset/7009 |
| Census 2011: Houses, Household Amenities and Assets Households by Main Source of Lighting [HH-7] | 7010 | https://ndap.niti.gov.in/dataset/7010 |
| Census 2011: Houses, Household Amenities and Assets Households by Ownership Status of the Census Houses , Size of the Household and Number of Dwelling Rooms [HH-4] | 7007 | https://ndap.niti.gov.in/dataset/7007 |
| Census 2011: Houses, Household Amenities and Assets Households by Predominant Material of Floor of Census Houses Occupied by them [HH-2C] | 7005 | https://ndap.niti.gov.in/dataset/7005 |
| Census 2011: Houses, Household Amenities and Assets Households by Predominant Material of Roof of Census Houses Occupied by them [HH-2A] | 7003 | https://ndap.niti.gov.in/dataset/7003 |
| Census 2011: Houses, Household Amenities and Assets Households by Predominant Material of Wall of Census Houses Occupied by them [HH-2B] | 7004 | https://ndap.niti.gov.in/dataset/7004 |

| | | |
|---|---|---|
| Census 2011: Houses, Household Amenities and Assets Households by the Condition of Census Houses Occupied by them [HH-1] | 6999 | https://ndap.niti.gov.in/dataset/6999 |
| Census 2011: Houses, Household Amenities and Assets Number of Households Availing Banking Services and Number of Households Having Each of the Specified Assets [HH-12] | 7002 | https://ndap.niti.gov.in/dataset/7002 |
| National Family Health Survey - 4 : District | 7034 | https://ndap.niti.gov.in/dataset/7034 |

**Supplemental Table 2**: Domains and indicators for socioeconomic development in the Census of India 2011 and the National Family Health Survey-4

| Domains | Indicators | |
|---|---|---|
| | Census of India, 2011 | NFHS-4 |
| **Sociodemographic** | - Urban population<br>- Rural population<br>- Population between 0-6 years of age<br>- Scheduled caste or tribe population | - Population below 15 years of age<br>- Sex ratio at birth for children born in the last five years<br>- Women age group 20 to 24 years married before age 18 years |
| **Socioeconomic** | - Literate women<br>- Working women | - Men aged 15 to 49 years who are literate<br>- Women aged 15 to 49 years who are literate |
| **Housing** | - Houses with good living conditions<br>- Houses with roofs made of pucca material<br>- Houses with walls made of pucca material<br>- Houses with floors made of pucca material<br>- Ownership status of homes<br>- Drinking water from a treated tap water source<br>- Electricity as source of lighting<br>- Improved latrine facilities<br>- Bathing facilities on house premises<br>- Bathing facilities connected to a closed drainage<br>- Kitchen inside house premises<br>- LPG as cooking fuel<br>- Any of the specified assets<br>- Access to banking facilities | - Clean cooking fuel<br>- Improved drinking water source<br>- Improved sanitation facility<br>- Households with electricity |
| **Community infrastructure or resources** | • At the town level:<br>- No. of latrines (pit and flush and pour)<br>- No. of domestic electric connections<br>- No. of industrial electric connections<br>- No. of commercial electric connections<br>- No. of electric connections for road lighting<br>- No. of healthcare facilities<br>- No. of primary schools (government and private)<br>- No. of middle schools (government and private)<br>- No. of secondary schools (government and private)<br>- No. of senior secondary schools (government and private) | - Institutional births<br>- Births delivered by skilled healthcare personnel<br>- Registered pregnancies<br>- Children under age 5 years whose birth was registered with the civil authority<br>- Households with any usual member covered under insurance scheme<br>- Mothers who received financial assistance under janani suraksha yojana |

- At the village level:

- No. of primary schools (government and private)
- No. of middle schools (government and private)
- No. of secondary schools (government and private)
- No. of senior secondary schools (government and private)
- No. of healthcare facilities

**Supplemental Table 3**: Domains and indicators for socioeconomic deprivation in the Multidimensional Poverty Index for India

| Domain | Indicator |
|---|---|
| **Health** | - Nutrition<br>- Child and adolescent mortality<br>- Antenatal care |
| **Education** | - Years of schooling<br>- School attendance |
| **Standard of Living** | - Cooking fuel<br>- Sanitation<br>- Drinking water<br>- Electricity<br>- Housing<br>- Assets<br>- Bank account |

**Supplemental Table 4:** Population and community characteristics- National Family Health Survey-4, Census of India, by urban-rural areas

| Census of India 2011 | | |
|---|---|---|
| **Indicator** | **Urban areas** | **Rural areas** |
| No. of latrines (pit and flush and pour) | 3992 (2266-7062) | - |
| No. of domestic electric connections | 1703 (1291-2087) | - |
| No. of industrial electric connections | 26 (12-48) | - |
| No. of commercial electric connections | 263 (164-401) | - |
| No. of electric connections for road lighting | 250 (116-369) | - |
| No. of healthcare facilities | 2 (1-3) | 5 (3-7) |
| No. of primary schools (government and private) | 6 (4-8) | 11 (8-15) |
| No. of middle schools (government and private) | 3 (2-5) | 5 (3-7) |
| No. of secondary schools (government and private) | 2 (1-3) | 2 (1-3) |
| No. of senior secondary schools (government and private) | 1 (1-2) | 1 (0-1) |
| Houses with good living conditions | 27537 (9596-69774) | 83542 (42578-138512) |
| % houses with roofs made of pucca material | 94 (90-97) | 85 (71-93) |
| % houses with walls made of pucca material | 78 (66-87) | 50 (26-68) |
| % houses with floors made of pucca material | 79 (62-91) | 27 (13-58) |
| % houses owned at the district level | 74 (62-84) | 96 (91-98) |
| % houses with drinking water from a treated tap water source | 51 (30-68) | 14 (5-27) |
| % houses that use electricity as source of lighting | 93 (82-97) | 70 (35-89) |
| % houses with latrine facility on premises- with improved facilities | 74 (63-84) | 26 (14-45) |
| % houses with bathing facilities on house premises | 71 (54-83) | 20 (9-44) |
| % houses with bathing facilities connected to a closed drainage | 22 (13-37) | 4 (2-8) |
| % houses with a kitchen inside the premises | 76 (64-87) | 51 (34-74) |
| % houses that use LPG as cooking fuel | 58 (45-70) | 8 (3-19) |
| % houses with any of the specified assets | 91 (87-94) | 78 (68-87) |
| % houses with banking facilities | 65 (59-73) | 53 (41-68) |
| % of population between 0-6 years of age | 12 (11-13) | 14 (12-17) |
| % of scheduled caste or tribe population | 17 (12-24) | 29 (21-46) |
| % of literate women | 33 (30-36) | 24 (20-29) |
| % of working women | 7 (5-9) | 17 (10-22) |
| Population | 248462 (87625-541660) | 940922 (479076-1501872) |

| National Family Health Survey-4 | | |
| --- | --- | --- |
| **Indicator** | **Urban areas *** | **Rural areas** |
| Households using clean fuel for cooking (%) | 87 (77-92) | 17 (9-36) |
| Population living in households with an improved drinking-water source (%) | 96 (91-98) | 91 (76-97) |
| Population living in households that use an improved sanitation facility (%) | 72 (64-84) | 36 (20-57) |
| Population living in households with electricity (%) | 99 (99-100) | 92 (77-98) |
| Population below age 15 years (%) | 23 (21-25) | 29 (24-34) |
| Sex ratio at birth for children born in the last five years[1] | 881 (792-1036) | 922 (846-1008) |
| Women in the age group of 20 to 24 years married before age 18 years (%) | 13 (8-18) | 24 (14-37) |
| Men age group 15 to 49 years who are literate (%) | 42 (41-43) | 83 (76-90) |
| Women age group 15 to 49 years who are literate (%) | 84 (80-89) | 64 (51-74) |
| Institutional births (%) | 95 (90-98) | 79 (61-91) |
| Births delivered by skilled health personnel (%) | 96 (90-99) | 81 (66-92) |
| Registered pregnancies (%) | 91 (83-96) | 92 (84-96) |
| Children under age 5 years whose birth was registered with the civil authority (%) | 96 (93-99) | 82 (64-96) |
| Households with any usual member covered under insurance scheme (%) | 28 (17-46) | 19 (7-48) |
| Mothers who received financial assistance under janani suraksha yojana (%) | 17 (7-29) | 38 (19-64) |

Data are presented as median (interquartile range)
* Restricted to districts with an urban population of at least 50%
[1] Sex ratio calculated as the number of females per 1000 males

**Supplemental Table 5**: Comparison of factor loadings (≥ 0.20) of retained indicators from principal component analysis in the NFHS-4 and Census

| NFHS-4 | | |
|---|---|---|
| Overall score | Rural score | Urban score |
| First principal component explained **42%** of total variance | First principal component explained **51%** of total variance | First principal component explained **91%** of total variance |
| **Housing indicators**<br>- Clean cooking fuel<br>- Improved sanitation facility<br>- Households with electricity | **Housing indicators**<br>- Improved drinking water source<br>- Improved sanitation facility<br>- Households with electricity | **Housing indicators**<br>- Clean cooking fuel<br>- Improved drinking water source<br>- Improved sanitation facility<br>- Households with electricity |
| **Sociodemographic indicators**<br>- Population below 15 years of age<br>- Women age group 20 to 24 years married before age 18 years | **Sociodemographic indicators**<br>- Sex ratio at birth for children born in the last five years | **Sociodemographic indicators**<br>- Population below 15 years of age<br>- Sex ratio at birth for children born in the last five years<br>- Women age group 20 to 24 years married before age 18 years |
| **Socioeconomic indicators**<br>- Men 15 to 49 years who are literate<br>- Women 15 to 49 years who are literate | **Socioeconomic indicators**<br>- Men 15 to 49 years who are literate<br>- Women 15 to 49 years who are literate | **Socioeconomic indicators**<br>- Men 15 to 49 years who are literate<br>- Women 15 to 49 years who are literate |
| **Infrastructure/Community resources**<br>- Institutional births<br>- Births delivered by skilled healthcare personnel<br>- Children under age 5 years whose birth was registered with the civil authority<br>- Mothers who received financial assistance under janani suraksha yojana | **Infrastructure/Community resources**<br>- Institutional births<br>- Births delivered by skilled healthcare personnel<br>- Children under age 5 years whose birth was registered with the civil authority<br>- Registered pregnancies | **Infrastructure/Community resources**<br>- Institutional births<br>- Births delivered by skilled healthcare personnel<br>- Registered pregnancies<br>- Children under age 5 years whose birth was registered with the civil authority<br>- Households with any usual member covered under insurance scheme<br>- Mothers who received financial assistance under janani suraksha yojana |
| On excluding non-contributing indicators, the first principal component explained **55%** of total variance | On excluding non-contributing indicators, the first principal component explained **70%** of total variance | All indicators were retained for the urban score |

| Census | | |
|---|---|---|
| Overall score | Rural score | Urban score |
| First principal component explained **29%** of total variance<br>**Housing indicators**<br>- Houses with walls made of pucca material<br>- Houses with floors made of pucca material<br>- Drinking water from a treated tap water source<br>- Electricity as source of lighting<br>- Improved latrine facilities<br>- Bathing facilities on house premises<br>- Bathing facilities connected to a closed drainage<br>- LPG as cooking fuel<br>- Any of the specified assets<br><br>**Sociodemographic indicators**<br>- Population between 0-6 years of age<br>Socioeconomic indicators<br>- Literate women<br><br>**Infrastructure/Community resources**<br>- N/A<br>On excluding non-contributing indicators, the first principal component explained **65%** of total variance | First principal component explained **32%** of total variance<br>**Housing indicators**<br>- Houses with walls made of pucca material<br>- Houses with floors made of pucca material<br>- Drinking water from a treated tap water source<br>- Electricity as source of lighting<br>- Improved latrine facilities<br>- Bathing facilities on house premises<br>- Bathing facilities connected to a closed drainage<br>- LPG as cooking fuel<br>- Any of the specified assets<br>- Access to banking facilities<br>**Sociodemographic indicators**<br>- Population between 0-6 years of age<br>Socioeconomic indicators<br>- Literate women<br><br>**Infrastructure/Community resources**<br>- N/A<br>On excluding non-contributing indicators, the first principal component explained **54%** of total variance | First principal component explained **28%** of total variance<br>**Housing indicators**<br>- Houses with floors made of pucca material<br>- Drinking water from a treated tap water source<br>- Electricity as source of lighting<br>- Improved latrine facilities<br>- Bathing facilities on house premises<br>- Bathing facilities connected to a closed drainage<br>- Kitchen inside house premises<br>- LPG as cooking fuel<br>- Any of the specified assets<br><br>**Sociodemographic indicators**<br>- Population between 0-6 years of age<br>Socioeconomic indicators<br>- N/A<br><br>**Infrastructure/Community resources**<br>- No. of domestic electric connections<br>On excluding non-contributing indicators, the first principal component explained **57%** of total variance |

**Supplemental Table 6**: Correlations between district-level socioeconomic development rural scores and health outcomes in the National Family Health Survey-4

| Health outcomes | NFHS-4 score (PCA) | NFHS-4 score (percentile) | Census score (PCA) | Census score (percentile) |
|---|---|---|---|---|
| Children under age 3 years breastfed within one hour of birth (%) | 0.18 | 0.26 | 0.04 | 0.02 |
| Breastfeeding children age group 6 to 23 months receiving an adequate diet (%) | 0.10 | 0.17 | 0.10 | 0.07 |
| Children age group 6 to 23 months receiving an adequate diet (%) | 0.13 | 0.22 | 0.17 | 0.13 |
| Children under 5 years who are stunted ( height-for-age ) (%) | -0.27 | -0.37 | **-0.63** | **-0.58** |
| Children under 5 years who are underweight ( weight-for-age ) (%) | -0.16 | -0.28 | **-0.52** | -0.49 |
| Children under 5 years who are wasted ( weight-for-height ) (%) | -0.01 | -0.08 | -0.20 | -0.16 |
| Children under 5 years who are severely wasted ( weight-for-height ) (%) | 0.00 | -0.04 | -0.13 | -0.09 |
| Children age group 6 to 59 months who are anemic (%) | -0.08 | -0.15 | -0.06 | -0.07 |
| Non-pregnant women age group 15 to 49 years who are anemic (%) | -0.02 | -0.06 | -0.09 | -0.16 |
| Women age group 15 to 49 years who are anemic (%) | -0.02 | -0.06 | -0.10 | -0.16 |
| Men age group 15 to 49 years who are anemic (%) | -0.04 | -0.08 | -0.32 | -0.35 |
| Women with body mass index below normal (%) | -0.09 | -0.22 | **-0.50** | -0.45 |
| Men with body mass index below normal (%) | -0.10 | -0.21 | -0.42 | -0.39 |
| Women who are overweight or obese (%) | 0.10 | 0.24 | **0.78** | **0.65** |
| Men who are overweight or obese (%) | 0.14 | 0.27 | **0.68** | **0.58** |
| Women suffering from very high blood sugar level (%) | 0.09 | 0.20 | 0.44 | 0.29 |
| Women suffering from high blood sugar level (%) | 0.08 | 0.17 | 0.34 | 0.21 |
| Men suffering from very high blood sugar level (%) | 0.07 | 0.14 | 0.17 | 0.06 |
| Men suffering from high blood sugar level (%) | 0.06 | 0.12 | 0.14 | 0.03 |
| Women with very high hypertension (%) | -0.02 | -0.02 | -0.05 | -0.02 |
| Women with moderately high hypertension (%) | 0.06 | 0.09 | 0.09 | 0.09 |
| Women-mildly elevated blood pressure (%) | 0.09 | 0.12 | 0.13 | 0.15 |
| Men with very high hypertension | 0.04 | 0.06 | 0.08 | 0.09 |
| Men with moderately high hypertension (%) | 0.06 | 0.08 | 0.20 | 0.23 |
| Men-mildly elevated blood pressure (%) | 0.17 | 0.21 | 0.29 | 0.31 |

**Supplemental Table 7**: Correlations between district-level socioeconomic development urban scores and health outcomes in the National Family Health Survey-4

| Health outcomes | NFHS-4 score (PCA) | NFHS-4 score (percentile) | Census score (PCA) | Census score (percentile) |
|---|---|---|---|---|
| Children under age 3 years breastfed within one hour of birth (%) | 0.10 | 0.03 | 0.05 | 0.07 |
| Breastfeeding children age group 6 to 23 months receiving an adequate diet (%) | 0.20 | 0.13 | -0.09 | -0.13 |
| Children age group 6 to 23 months receiving an adequate diet (%) | 0.23 | 0.16 | -0.08 | -0.15 |
| Children under 5 years who are stunted ( height-for-age ) (%) | -0.15 | -0.20 | -0.23 | -0.20 |
| Children under 5 years who are underweight ( weight-for-age ) (%) | -0.03 | -0.02 | -0.12 | -0.17 |
| Children under 5 years who are wasted ( weight-for-height ) (%) | 0.04 | 0.05 | 0.09 | -0.03 |
| Children under 5 years who are severely wasted ( weight-for-height ) (%) | 0.01 | -0.00 | 0.08 | -0.02 |
| Children age group 6 to 59 months who are anemic (%) | -0.12 | -0.06 | 0.03 | -0.13 |
| Non-pregnant women age group 15 to 49 years who are anemic (%) | 0.04 | 0.06 | -0.08 | -0.26 |
| Women age group 15 to 49 years who are anemic (%) | 0.04 | 0.07 | -0.08 | -0.26 |
| Men age group 15 to 49 years who are anemic (%) | 0.08 | 0.12 | -0.22 | -0.26 |
| Women with body mass index below normal (%) | 0.03 | 0.03 | -0.27 | -0.22 |
| Men with body mass index below normal (%) | -0.00 | -0.01 | -0.34 | -0.33 |
| Women who are overweight or obese (%) | -0.01 | 0.07 | 0.50 | 0.35 |
| Men who are overweight or obese (%) | 0.08 | 0.14 | 0.58 | 0.46 |
| Women suffering from very high blood sugar level (%) | 0.17 | 0.14 | -0.01 | -0.01 |
| Women suffering from high blood sugar level (%) | 0.13 | 0.09 | -0.11 | -0.07 |
| Men suffering from very high blood sugar level (%) | 0.13 | 0.12 | 0.06 | 0.10 |
| Men suffering from high blood sugar level (%) | 0.17 | 0.15 | -0.01 | 0.06 |
| Women with very high hypertension (%) | **-0.69** | **-0.78** | -0.22 | -0.17 |
| Women with moderately high hypertension (%) | -0.34 | -0.38 | -0.08 | -0.10 |
| Women-mildly elevated blood pressure (%) | -0.46 | -0.44 | 0.03 | 0.03 |
| Men with very high hypertension | -0.10 | -0.08 | 0.20 | 0.19 |
| Men with moderately high hypertension (%) | 0.06 | 0.05 | 0.14 | 0.16 |
| Men-mildly elevated blood pressure (%) | -0.30 | -0.27 | 0.08 | 0.06 |

* Urban areas in NFHS-4 restricted to districts with at least 50% of urban population
